# Genome-wide determinants of mortality and motor progression in Parkinson’s disease

**DOI:** 10.1101/2022.07.07.22277297

**Authors:** Manuela MX Tan, Michael A Lawton, Miriam I Pollard, Emmeline Brown, Raquel Real, Alejandro Martinez Carrasco, Samir Bekadar, Edwin Jabbari, Regina H Reynolds, Hirotaka Iwaki, Cornelis Blauwendraat, Sofia Kanavou, Leon Hubbard, Naveed Malek, Katherine A Grosset, Nin Bajaj, Roger A Barker, David J Burn, Catherine Bresner, Thomas Foltynie, Nicholas W Wood, Caroline H Williams-Gray, Ole A Andreassen, Mathias Toft, Alexis Elbaz, Fanny Artaud, Alexis Brice, Jean-Christophe Corvol, Jan Aasly, Matthew J Farrer, Michael A Nalls, Andrew B Singleton, Nigel M Williams, Yoav Ben-Shlomo, John Hardy, Michele TM Hu, Donald G Grosset, Maryam Shoai, Lasse Pihlstrøm, Huw R Morris

## Abstract

**Background:** There are 90 genetic risk variants for Parkinson’s disease (PD) but currently only five nominated loci for PD progression. The biology of PD progression is likely to be of central importance in defining mechanisms that can be used to develop new treatments.

**Methods:** We studied 6,766 PD patients, over 15,340 visits with a mean follow-up of between 4.2 and 15.7 years and carried out a genome wide survival study for time to a motor progression endpoint, defined by reaching Hoehn and Yahr stage 3 or greater, and death (mortality).

**Findings:** There was a robust effect of the ***APOE*** ε4 allele on mortality in PD. We identified three novel loci for mortality and motor progression, and nominated genes based on physical proximity and/or expression quantitative trait loci data. One locus within the ***TBXAS1*** gene encoding thromboxane A synthase 1 was associated with mortality in PD (HR = 2.04 [95% CI 1.63 to 2.56], p-value = 7.71 x 10^-10^). Another locus near the ***SYT10*** gene encoding synaptotagmin 10 was associated with mortality just above genome-wide significance (HR=1.36 [95% CI 1.21 to 1.51], p-value=5.31×10^-8^). We also report 4 independent loci associated with motor progression: the top locus within ***MORN1*** (HR=2.76 [95% CI 1.97 to 3.87], p-value=3.1×10^-9^), the second most significant locus near ***ASNS***, the third most significant locus near ***PDE5A***, and a fourth locus within ***XPO1***. We have nominated causal genes based on physical position, however we also discuss other possible causal genes based on expression quantitative trait loci, colocalization analysis, and tagging of rare variants. Only the non-Gaucher disease causing ***GBA1*** PD risk variant E326K, of the known PD risk variants, was associated with mortality in PD.

**Interpretation:** We report six novel loci associated with PD motor progression or mortality. Further work is needed to understand the links between these genomic variants and the underlying disease biology. However, thromboxane synthesis, vesicular peptidergic neurotransmitter release, and phosphodiesterase inhibition may represent new candidates for disease modification in PD.

**Funding sources:** Parkinson’s UK, Aligning Science Across Parkinson’s through the Michael J Fox Foundation for Parkinson’s Research, Southern and Eastern Norway Regional Health Authority

## INTRODUCTION

Parkinson’s disease (PD) is a progressive neurodegenerative condition for which there are no treatments to stop or slow disease progression. Large-scale genome-wide case-control association studies (GWASs) of PD have identified 90 independent variants associated with disease risk^1^. However, it is also important to study the genetics and biology of disease progression. This will enable the development of potential disease-modifying treatments. There have now been a handful of GWAS which aim to identify genetic variants associated with progression in PD. These have nominated loci in *SLC44A1* (encoding choline transporter like protein -1, involved in membrane synthesis) for progression to Hoehn and Yahr (H&Y) stage 3 or greater,^2^ *APOE* for cognitive progression,^3^ *LRP1B* (encoding an low-density lipoprotein receptor which is involved in amyloid precursor protein trafficking) for progression to dementia^4^, and *RIMS2* (encoding the RAB3 interacting RIMS2 protein, involved in neurotransmitter release) for progression to PD dementia.^5^ In addition, many candidate gene studies have shown that variants in *GBA1*, *APOE,* and potentially *MAPT*, are associated with the rate of PD motor and cognitive progression.^6^

PD progression may be determined by differential cellular susceptibility, related to mitochondrial function or proteostasis, differential cell to cell spread of pathology, or novel pathways and mechanisms. Risk factors determined from case control studies indicate aetiological pathways and guide future preventive trials, but these may differ from risk factors that determine disease progression. Currently disease modifying treatment trials focus on intervention in recently diagnosed patients, related to disease progression after diagnosis. Work on large scale longitudinal cohorts over the last ten years has now enabled the collaborative study of large clinico-genetic datasets. Here we have carried out two progression GWASs: progression to mortality, and Hoehn and Yahr (H&Y) stage 3 or greater. We have analysed data from 6,766 PD patients with over 15,340 visits and mean follow-up ranging between 4.2 to 15.7 years.

## METHODS

We studied 11 cohorts from Europe and America, and included cohorts in each analysis who had sufficient data on the outcomes of interest (see <u>Supplementary Materials</u>). Genotyping, quality control, and imputation was performed in each cohort separately but following the same steps. Only variants with high imputation quality scores (INFO/R2 > 0.8) and minor allele frequency (MAF) > 1% were retained for analysis.

We assessed the following two clinical outcomes: mortality, and H&Y stage 3 or greater. Cohorts were excluded if less than 20 individuals met the outcome of interest during the follow-up period, or < 5% of the total cohort size. Progression to the clinical milestones was analysed using Cox proportional hazard models. Progression to mortality was assessed from the starting timepoint of PD onset. Progression to Hoehn and Yahr stage 3 or greater was assessed from the time from study entry (baseline visit). Individuals who met the outcome of Hoehn and Yahr stage 3 or greater at study entry were left censored and excluded from analyses. In each model, we adjusted for age at onset, gender, and the first five genetic principal components to adjust for population stratification. Meta-analysis was performed in METAL (version 2011-03-25)^7^, using an inverse variance weighted fixed effects model. GWASs with a genomic inflation factor above 1.2 were excluded from the meta-analysis. Only SNPs that were genotyped in > 1,000 individuals across all cohorts were included in the final results. SNPs with heterogeneous effects across cohorts were also excluded (p-value < 0.05 for Cochran’s Q-test for heterogeneity, and/or I^2^ > 80). The null hypothesis was tested with the standard GWAS significance level of 5 x 10^-8^. Results were uploaded to Functional Mapping and Annotation of GWAS (FUMA; https://fuma.ctglab.nl/)^8^ to annotate, prioritise, and visualize GWAS results, and perform gene-set analysis with MAGMA. Genome-wide Complex Trait Analysis conditional and joint analysis (GCTA-COJO, version 1.94.1) was used to identify independently-associated SNPs^9^, using all individuals in the Accelerating Medicines Partnership Parkinson’s disease (AMP-PD) whole genome sequencing dataset as a reference sample (N = 9422). Fine-mapping of the top loci was performed with Probabilistic Annotation INtegraTOR (PAINTOR) (https://bogdan.dgsom.ucla.edu/pages/paintor/) following the recommended pipeline, to prioritize causal variants and integrate functional genomic data^10–12^. Forest plots for the top SNPs were generated in R v3.6 using the *forestplot* package, enabling us to determine consistency, and replication of survival risk alleles across cohorts. Further details are provided in the <u>Supplementary Materials</u>.

We also performed candidate variant analysis of the 90 PD risk variants from the most recent PD case-control GWAS,^1^ and the cumulative PD genetic risk score (GRS). We also examined associations for other candidate variants that have been reported in PD and Progressive Supranuclear Palsy (PSP) progression: *SLC44A1*^2^*, RIMS2*, *WWOX, TMEM108*^5^, *APOE* ε2 allele, *MAPT* H1 haplotype, and rs2242367 adjacent to the *LRRK2* locus^13^. We analysed the Alzheimer’s disease (AD) GRS in relation to PD progression. 38 loci passing genome-wide significance from the latest AD GWAS were used to create the AD GRS.^14^ The *APOE* region was excluded from the GRS (hg19/GRCh37 coordinates 19:40,000,000-50,000,000).^14^

## RESULTS

Overall 6,766 participants with PD were analysed with mean follow-up between 4.2 and 15.7 years (Table 1). We did not have data from regular follow-up visits for all studies as some studies only contributed mortality data. However, in the studies that had regular follow-up visit data available, over 15,340 visits were analysed (Table 1).

**Table 1.**
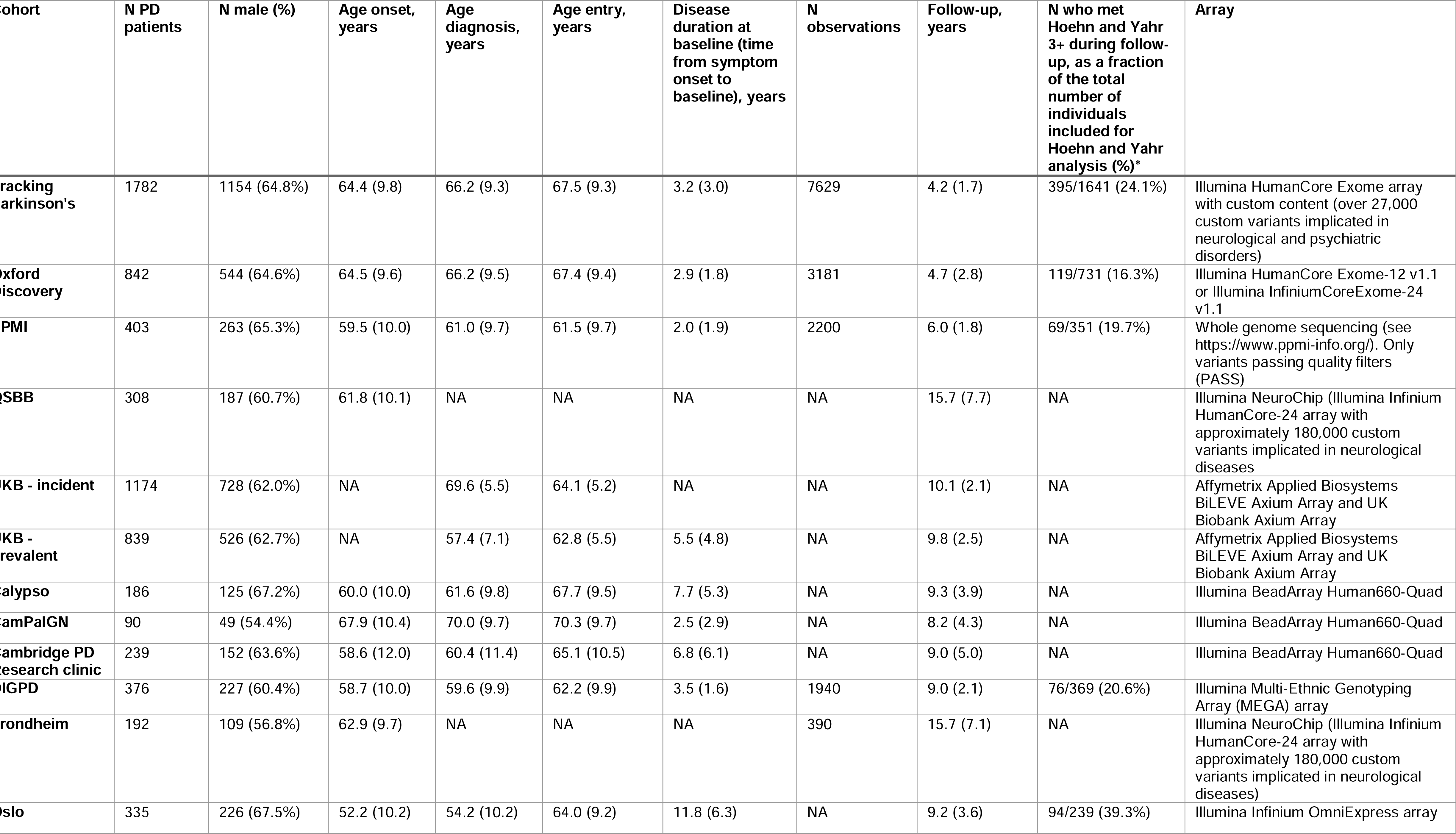
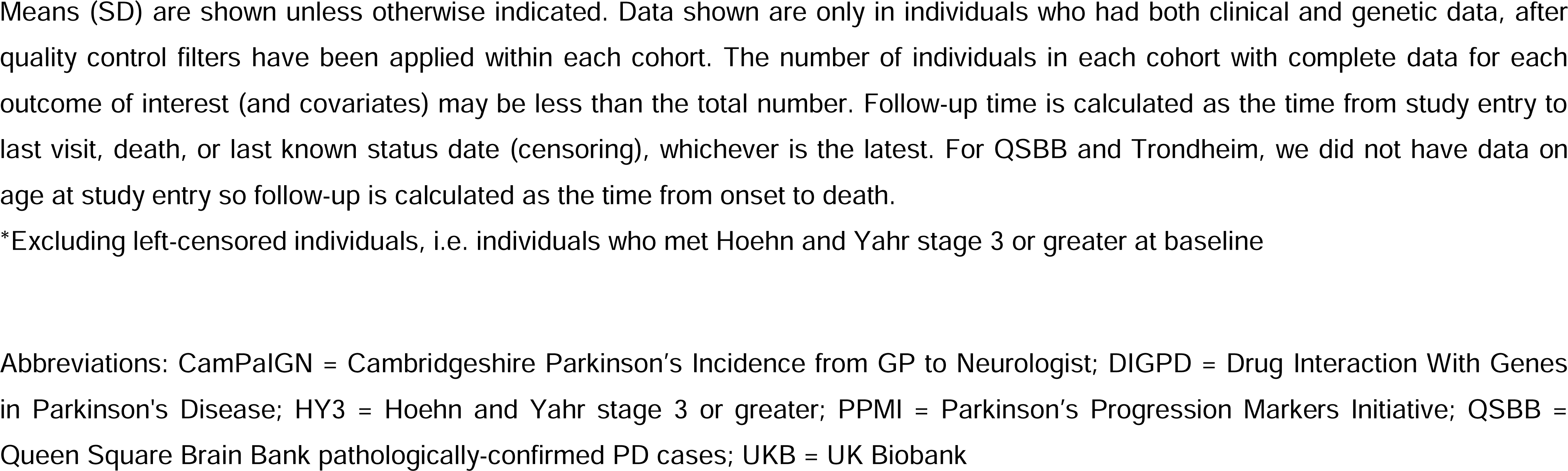
Cohort demographics.

### GWAS of mortality

One study was excluded from the meta-analysis of mortality because the study-specific genomic inflation factor was above 1.2 and one study was excluded because less than 20 individuals reached this endpoint (Supplementary Table 1).

5,744 patients were included in the meta-analysis of mortality. Of these, 1,846 (32.1%) individuals had died with a median time to death of 10.6 years from PD onset. 7,696,389 SNPs were present in at least 1,000 individuals and 7,313,918 SNPs passed meta-analysis filtering for heterogeneity and MAF variability. The genomic inflation value of the meta-analysis after filtering was 1.04.

Two loci passed genome-wide significance and were identified to determine mortality in PD (Figure 1). The top SNP was rs429358 in Chromosome 19 (p=4.0×10^-10^), which tags the *APOE* ε4 allele (Table 2). One other locus in Chromosome 7 in *TBXAS1* also reached significance (p=7.7×10^-10^), and another locus in Chromosome 12 near *SYT10* was nominally associated (p=5.3×10^-8^). Regional association plots are shown in Figure 1 and Supplementary Figures 1-3. The top ten independent SNPs identified from GCTA-COJO and their nearest genes are reported in Table 2.

**Figure 1.**
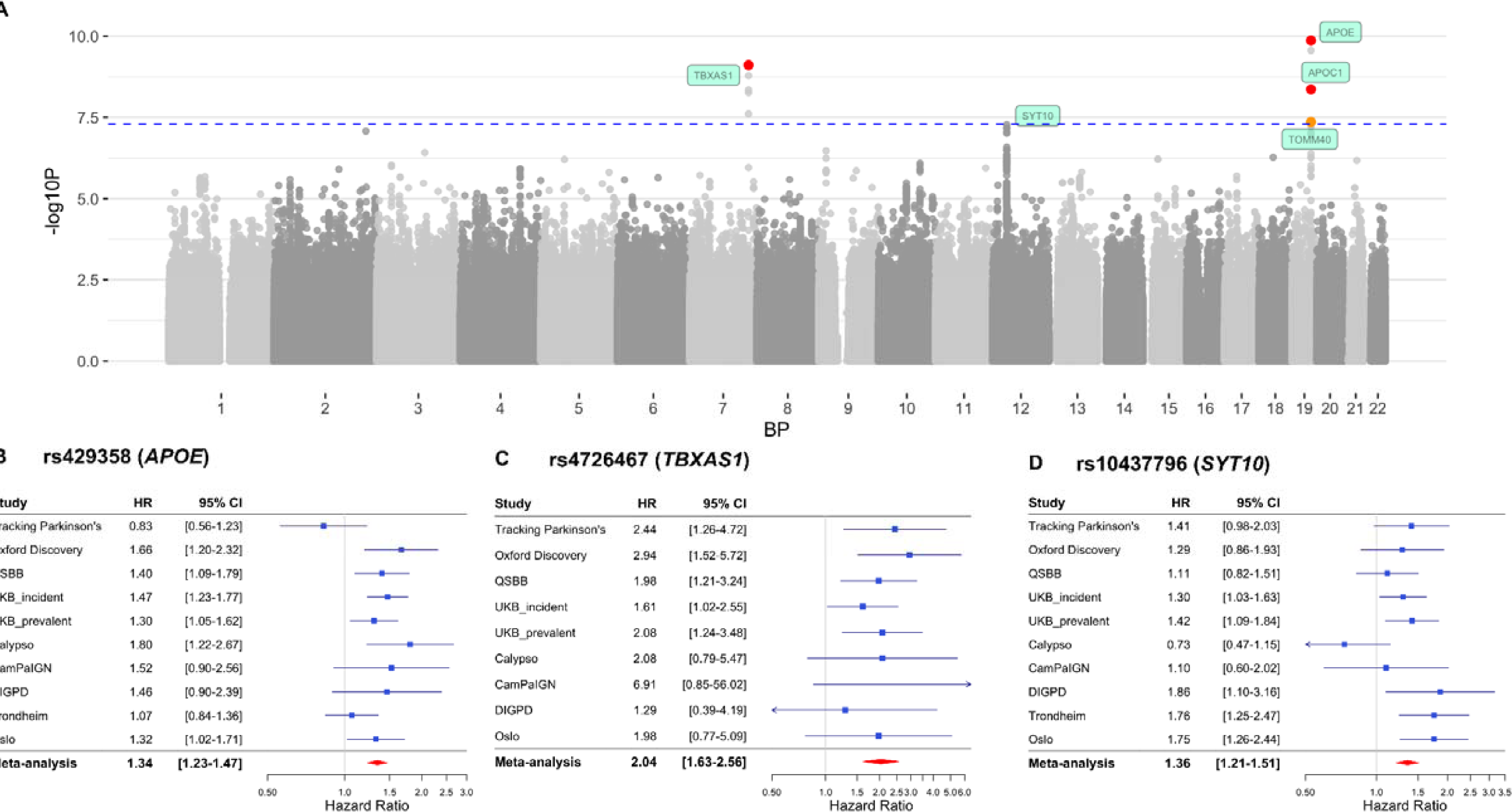
GWAS meta-analysis of mortality. **(A)** The Manhattan plot showing two GWAS significant loci after meta-analysis. The blue dashed line indicates the threshold for genome-wide significance, p=5×10^-8^. SNPs highlighted in red have p-value<5×10^-9^. SNPs highlighted in orange have p-value<5×10^-8^. One nominal association in Chromosome 12 is also annotated with the nearest gene, *SYT10* (p=5.3×10^-8^). **(B)** Forest plot for the top SNP rs429358 in Chromosome 19, in *APOE*. **(C)** Forest plot for the top SNP rs4726467 in Chromosome 7, in *TBXAS1*. **(D)** Forest plot for the top SNP rs10437796 in Chromosome 12, near *SYT10.* BP *=* Base Pair, CI = Confidence Interval, HR = Hazard Ratio.

**Table 2.** Top 10 independent SNPs from meta-analysis of progression to mortality.

| chr | bp | rsID | effect allele | non-effect allele | effect allele freq | nearest gene | distance to gene (BP) | beta | SE | Hazard Ratio | 95% CI | p-value | p-value COJO |
| --- | --- | --- | --- | --- | --- | --- | --- | --- | --- | --- | --- | --- | --- |
| **19** | 45411941 | rs429358 | C | T | 0.160 | *APOE* | 0 | 0.295 | 0.046 | 1.342 | 1.23-1.47 | 1.35E-10 | 1.60E-10 |
| **7** | 139637422 | rs4726467 | T | C | 0.021 | *TBXAS1* | 0 | 0.713 | 0.116 | 2.039 | 1.63-2.56 | 7.71E-10 | 8.49E-10 |
| **12** | 33635494 | rs10437796 | A | C | 0.098 | *SYT10* | 42740 | 0.304 | 0.056 | 1.355 | 1.21-1.51 | 5.31E-08 | 5.76E-08 |
| **9** | 17616880 | rs3808753 | G | A | 0.035 | *SH3GL2* | 0 | 0.448 | 0.088 | 1.564 | 1.32-1.86 | 3.34E-07 | 3.52E-07 |
| **3** | 113979619 | rs142285045 | A | C | 0.013 | *ZNF80* | 23194 | 0.836 | 0.165 | 2.308 | 1.67-3.19 | 3.80E-07 | 4.06E-07 |
| **18** | 33965217 | rs76125680 | C | G | 0.012 | *FHOD3* | 0 | 0.874 | 0.174 | 2.397 | 1.70-3.37 | 5.32E-07 | 5.65E-07 |
| **15** | 34896795 | rs35294489 | G | A | 0.014 | *GOLGA8B* | 20959 | 0.757 | 0.152 | 2.131 | 1.58-2.87 | 6.07E-07 | 6.43E-07 |
| **5** | 55533638 | rs28811891 | T | C | 0.026 | *ANKRD55* | 4452 | 0.544 | 0.109 | 1.722 | 1.39-2.13 | 6.21E-07 | 6.49E-07 |
| **21** | 31807104 | rs145506557 | C | A | 0.016 | *KRTAP13-4* | 4028 | 0.896 | 0.180 | 2.451 | 1.72-3.49 | 6.59E-07 | 7.27E-07 |
| **10** | 98786658 | rs17112311 | A | G | 0.049 | *C10orf12* | 41073 | 0.364 | 0.074 | 1.439 | 1.25-1.66 | 8.07E-07 | 8.31E-07 |

Independent SNPs identified with GCTA-COJO.
Genome coordinates are in build hg19/GRCh37
Abbreviations: BP = base pair, chr = chromosome, CI = Confidence Interval, freq = frequency, GCTA-COJO = Genome-wide Complex Trait Analysis conditional and joint analysis, SE = Standard Error, SNP = Single Nucleotide Polymorphism

In the MAGMA gene-based test, *APOE* was significantly associated with mortality (p=1.9×10^-10^), and *SYT10* was associated just below genome-wide significance (p=3.6×10^-6^). There was no significant association of any gene-sets or tissues in the MAGMA gene-set analysis and gene property analysis for tissue specificity.

A locus in *TBXAS1* was also significantly associated with PD mortality, with the top SNP rs4726467. This SNP is an expression quantitative trait locus (eQTL), with the effect (minor) allele decreasing expression of *TBXAS1* in blood (eQTLGen; https://www.eqtlgen.org/)^15^ but not other tissues as reported in GTEx (https://gtexportal.org/). There was no evidence on GTEx that rs4726467 is a splicing Quantitative Trait Loci (sQTLs). Brain eQTL data at MetaBrain (https://www.metabrain.nl/)^16^ did not indicate this SNP was a significant cis-eQTL in any brain regions from European samples.

In LDproxy (https://ldlink.nci.nih.gov/), we identified two coding variants in linkage disequilibrium (LD) within 500kb of the top SNP. One was a synonymous variant in *HIPK2*, rs34093649 (D’ = 0.24, R^2^ = 0.05, MAF 0.02). There was also one missense variant in *PARP12,* rs2286196, about 89kb away from rs4726467 (D’ = 0.34, R^2^ = 0.01, MAF = 0.20).

The top SNP in Chromosome 12, rs10437796, is not directly within *SYT10* but increases *SYT10* expression in the testis and decreases expression in the tibial nerve. The SNP also increases expression of the long noncoding RNA (lncRNA) RP11-438D14.2 (ENSG00000259937) in the brain. This is a ‘sense intronic’ transcript to *SYT10*, a long non-coding transcript which is within an intron of a coding gene and does not overlap any exons. Brain cis-eQTL data from MetaBrain showed that the effect allele A of rs10437796 significantly increased expression of *SYT10* in the cortex but not in other brain regions. This SNP was not an eQTL or sQTL for any genes in blood in eQTLGen. There were no coding variants in LD with this SNP in LDproxy within a 500kb window.

We also performed colocalization analysis to determine whether the association signals for PD mortality and gene expression are driven by a shared causal variant (see <u>Supplementary Materials</u>; Supplementary Table 2). We used cis-eQTL data from PsychENCODE and eQTLGen to examine gene expression in whole brain or blood, respectively. However, no PD mortality loci showed evidence of colocalization with eQTLs (PP.H4 <0.75). The largest posterior probability was 0.42 for eQTLs regulating CLASRP in brain from the PsychENCODE database (Supplementary Table 2).

We checked the top SNPs and genes (+/- 1 Mb) from the mortality GWAS in the most recent longevity GWAS^17^ to see if these were influencing PD-specific or more general mortality and survival. *APOE* and specifically the ε4 tagging SNP, rs429358, was the strongest signal for longevity (beta = -0.05, p = 9.6×10^-127^). The *TBXAS1* SNP, rs4726467, was not associated with longevity (p = 0.82). Other SNPs within or just outside the *TBXAS1* gene boundaries were also not associated with longevity at genome-wide significance. The nearest associated SNP was rs149577943 which lay approximately 1Mb outside of *TBXAS1* (p = 6.1×10^-5^). The *SYT10* SNP, rs10437796, was nominally associated with reduced longevity (beta = -0.008, p=0.003) in the longevity GWAS.

### GWAS of H&Y stage 3 or greater

3,331 individuals across 5 cohorts were analysed for progression to H&Y stage 3 or greater. 753 individuals (22.6%) met the outcome of H&Y stage 3 or greater, with a median time to H&Y3 of 3.1 years. 6,549,622 SNPs passed filtering for heterogeneity and MAF variability. The genomic inflation factor of the meta-analysis was 1.03. The top ten independent SNPs from GCTA-COJO are reported in Table 3.

**Table 3.** Top 10 independent SNPs from meta-analysis of progression to Hoehn and Yahr stage 3 or greater.

| chr | bp | rsID | effect allele | non-effect allele | effect allele freq | nearest gene | distance to gene (BP) | beta | SE | Hazard Ratio | 95% CI | p-value | p-value COJO |
| --- | --- | --- | --- | --- | --- | --- | --- | --- | --- | --- | --- | --- | --- |
| 1 | 2315032 | rs115217673 | A | G | 0.016 | *MORN1* | 0 | 1.016 | 0.172 | 2.762 | 1.97-3.87 | 3.09E-09 | 3.53E-09 |
| 7 | 97470925 | rs145274312 | A | G | 0.011 | *ASNS* | 10504 | 1.872 | 0.317 | 6.503 | 3.49-12.11 | 3.49E-09 | NA* |
| 4 | 120566153 | rs113120976 | T | C | 0.013 | *PDE5A* | 16172 | 1.581 | 0.273 | 4.859 | 2.85-8.30 | 6.95E-09 | 8.72E-09 |
| 2 | 61742356 | rs141421624 | G | A | 0.017 | *XPO1* | 0 | 0.923 | 0.167 | 2.517 | 1.82-3.49 | 3.08E-08 | 3.36E-08 |
| 1 | 231267735 | rs74796254 | T | C | 0.027 | *LOC149373* | 52110 | 0.693 | 0.131 | 2.000 | 1.55-2.58 | 1.17E-07 | 1.27E-07 |
| 1 | 247425485 | rs72771919 | G | A | 0.021 | *VN1R5* | 5038 | 0.770 | 0.146 | 2.160 | 1.62-2.88 | 1.44E-07 | 1.55E-07 |
| 4 | 186697549 | rs75614365 | C | G | 0.121 | *SORBS2* | 0 | 0.372 | 0.071 | 1.450 | 1.26-1.67 | 1.94E-07 | 2.13E-07 |
| 1 | 43105085 | rs75140767 | G | A | 0.028 | *PPIH* | 18621 | 0.715 | 0.140 | 2.044 | 1.55-2.69 | 3.26E-07 | 3.49E-07 |
| 2 | 131557187 | rs7566590 | A | G | 0.042 | *AMER3* | 31480 | 0.573 | 0.112 | 1.774 | 1.42-2.21 | 3.31E-07 | 3.58E-07 |
| 6 | 11284774 | rs148949229 | A | G | 0.014 | *NEDD9* | 0 | 1.694 | 0.334 | 5.441 | 2.83-10.46 | 3.78E-07 | 4.72E-07 |
| 15 | 82198898 | rs58698247 | C | T | 0.024 | *MEX3B* | 135221 | 0.692 | 0.136 | 1.997 | 1.53-2.61 | 3.94E-07 | 4.20E-07 |

Independent SNPs identified with GCTA-COJO.
Genome coordinates are in build hg19/GRCh37
Abbreviations: BP = base pair, chr = chromosome, CI = Confidence Interval, freq = frequency, GCTA-COJO = Genome-wide Complex Trait Analysis conditional and joint analysis, SE = Standard Error, SNP = Single Nucleotide Polymorphism
\*One SNP, rs145274312, was not included in the COJO analysis as it was not present in the AMP-PD reference dataset. This is likely because the minor allele frequency is close to 1% in the general population (0.97% in gnomAD non-Finnish European population), however in our PD datasets the allele frequency was >1%

Four independent loci were significantly associated with progression to H&Y 3 or greater (p < 5 x 10^-8^). Regional association plots for all GWAS significant loci are shown in Supplementary Figures 4-7. The top locus was in Chromosome 1, with lead SNP rs115217673 within the *MORN1* gene (Figure 2). The top SNP was not a significant eQTL for any genes, according to GTEx, eQTLGen, or MetaBrain databases. As the *GBA1* gene is also located in Chromosome 1 and is an important factor for PD risk and progression, located 152 Mb away^18,19^, we checked whether the top SNP rs115217673 was in linkage disequilibrium with any of the common *GBA1* variants (p.E326K, p.N370S, p.T369M, and p.L444P) using the LDpair tool (https://ldlink.nih.gov/?tab=ldpair). However, there was no evidence of linkage disequilibrium between our top H&Y3 locus and any of the main *GBA1* variants (Supplementary Table 3).

**Figure 2.**
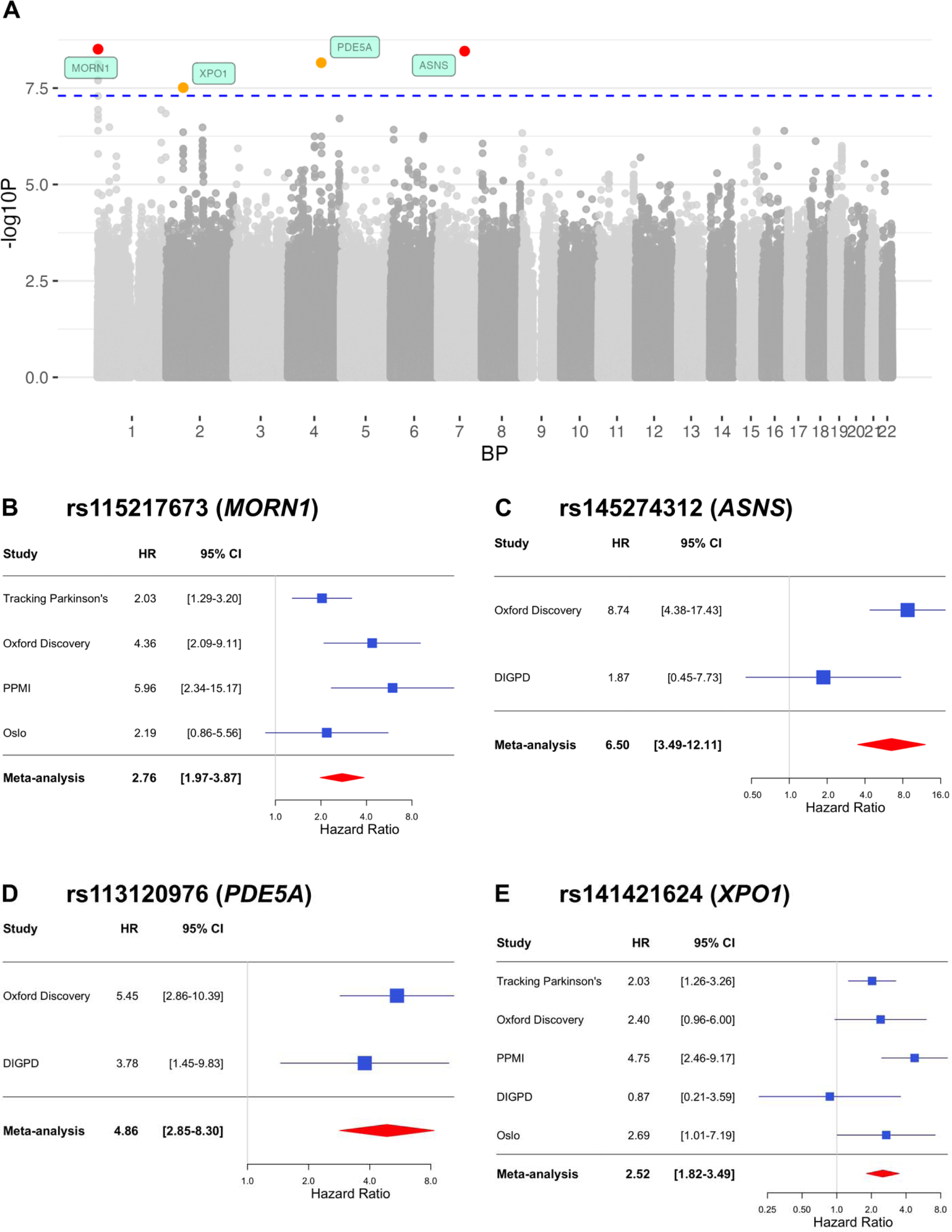
GWAS meta-analysis of progression to Hoehn and Yahr stage 3 or greater (H&Y3+). Note that the loci in the Manhattan plot and the forest plots have been labelled with the physically closest genes, though these may not necessarily be the causal genes. The Manhattan plot showing four GWAS significant loci after meta-analysis. The blue dashed line indicates the threshold for genome-wide significance, p=5×10^-8^. SNPs highlighted in red have p-value<5×10^-9^. SNPs highlighted in orange have p-value<5×10^-8^. Forest plot for the top SNP rs115217673 in Chromosome 1, in *MORN1*. **(C)** Forest plot for the top SNP rs145274312 in Chromosome 7, near *ASNS*. **(D)** Forest plot for the top SNP rs113120976 in Chromosome 4, near *PDE5A.* **(E)** Forest plot for the top SNP rs141421624 in Chromosome 2, in *XPO1.* BP *=* Base Pair, CI = Confidence Interval, HR = Hazard Ratio.

Published summary statistics from a previous progression GWAS^12^ were downloaded from https://pdgenetics.shinyapps.io/pdprogmetagwasbrowser/ (accessed November 2019). These summary statistics only included variants that passed heterogeneity filters, with MAF > 0.05 and total number of participants > 1000. Our top SNP in *MORN1* was not associated with progression to H&Y3 in the Iwaki summary statistics (beta = 0.61, p = 0.02).

The second most significant locus was in Chromosome 7, with top SNP rs145274312, nearest to the *ASNS* gene. This SNP was also not identified as an eQTL for any genes in the eQTL databases. This SNP was not included in any of the Iwaki summary statistics for H&Y stage, UPDRS2, or UPDRS3.

The third most significant locus was in Chromosome 4, with top SNP rs113120976. This was closest to the protein-coding gene *PDE5A,* however it is also near a long non-coding RNA *LOC107986192* (NR_165235.1). The top SNP is a significant cis-eQTL in blood for *USP53* with the effect allele T decreasing gene expression (eQTLGen). However, the SNP was not an eQTL in other tissues as reported in GTEx and MetaBrain. This SNP was also not included in any of the Iwaki GWAS summary statistics related to motor progression.

Finally, there was a locus in Chromosome 2 which was significantly associated with progression to H&Y3, with top SNP rs141421624. This SNP is intronic in the *XPO1* gene. However, it is also a significant eQTL in blood for the genes *KIAA1841* and *AHSA2* (eQTLGen). It is also a significant eQTL in brain cortex for *C2orf74,* with the effect allele G increasing gene expression (MetaBrain).

In LDproxy (https://analysistools.cancer.gov/LDlink), there were also two missense variants within 500kb in linkage disequilibrium with the lead SNP rs141421624. One of these is rs1729674, a missense variant in *C2orf74*, with a D’ of 1.0 and R^2^ of 0.014, and MAF of 0.42. The other variant in LD with the lead SNP is rs76248080, a missense variant in the *CCT4*, D’ = 0.55. R^2^ = 0.014, MAF = 0.18. This SNP was also not included in any of the Iwaki GWAS summary statistics related to motor progression.

Colocalization analysis did not provide strong evidence for shared causal variants between PD H&Y3 loci and gene expression, with no loci surpassing PP.H4 > 0.75. However, the strongest signal (highest PP.H4) was for the locus in Chromosome 2, which showed some evidence of colocalization with eQTLs for *PUS10* (PP.H4 = 0.69) at the liberal p12 of 1e^-5^. p12 is the prior probability that any random SNP in the region is associated with both traits, and though 1e-5 is the default in coloc, it may be too liberal particularly in smaller samples. Full results and figures of colocalization analysis are shown in Supplementary Table 4 and Supplementary Figure 8. There was no strong evidence of colocalization with eQTLs for any of the other PD H&Y3 loci.

### Candidate variant analysis

We did not find that any of the 90 PD risk SNPs were associated with PD progression at genome-wide significance (<u>Supplementary Materials</u>; Supplementary Table 5). Two SNPs, representing rare *LRRK2* (rs34637584) and *GBA1* (rs114138760) variants, were not present in our analysis as we filtered out variants with MAF < 1%. Of the 88 PD risk variants tested, we found that only one variant, rs35749011, near *KRTCAP2* and tagging the *GBA1* p.E326K variant, was significantly associated with mortality (p=3.6×10^-4^) below the analysis-wide significance threshold (p-value threshold 0.05/88=0.00057). There was also one other variant, rs62333164 in *CLCN3*, which was nominally associated with progression to H&Y3 (p=0.005) (Supplementary Figure 9).

We also examined the association between the PD GRS and progression in each cohort, adjusting for sex, age at onset, and PC1-PC5. The random-effects meta-analysis across cohorts did not show an effect of the GRS on mortality (HR = 0.99 [95% CI 0.95 to 1.04], p = 0.74), or H&Y3+ (HR = 1.02 [95% CI 0.96 to 1.08], p = 0.60).

In the candidate variant analysis, the PSP survival SNP rs2242367 was nominally associated with more rapid progression to mortality in PD (HR=1.13 [95% CI 1.04 to 1.21], p=0.002) (Supplementary Table 6).

### Alzheimer’s Disease Genetic Risk Score (GRS)

In the random-effects meta-analysis, the AD GRS without *APOE* was nominally associated with mortality (HR=1.06 [95% CI 1.01 to 1.11], p=0.03).

### Power calculations

The power to detect a signal in a survival GWAS depends on a number of factors, including effect size, allele frequency of the effect allele, and the proportion of individuals meeting the outcome of interest. Using the ‘survSNP’ package^20^, we estimate that this study had 92% power to detect a significant effect (p<5×10^-8^) for our top *APOE* SNP rs429358 in the mortality GWAS, given an allele frequency of 16%, Hazard Ratio of 1.34, event/death rate of 32.1% and median time to death of 10.6 years. Supplementary Figure 10 shows how power changes with different event rates and allele frequencies. Clearly power for progression studies will increase with longer follow-up as more individuals meet the outcomes.

## DISCUSSION

We have conducted a large meta-analysis GWAS of progression to clinical milestones in PD. We have identified the loci in or near *APOE, TBXAS1, MORN1, ASNS, PDE5A,* and *XPO1*, as relevant to survival and motor cognitive progression in PD. The effects were largely consistent and replicated across cohorts, as illustrated in the forest plots and by formal tests of heterogeneity.

### APOE

The *APOE* SNP rs429358 was associated with mortality. *APOE* is the strongest genetic risk factor for AD,^14,21^ and is also associated with cardiovascular disease (including coronary heart disease / coronary artery disease), and cholesterol levels.^22^

In PD, *APOE* has been associated with age at onset,^23^ cognition and dementia, and potentially motor progression,^24^ but not PD risk.^1,25^ In the GWAS of PD age at onset, the effect of *APOE* was shown to be similar between age at onset in cases and age of entry of controls.^23^ This suggests that the effect of *APOE* on PD age at onset is more generally related to ageing, and not specific to PD age at onset. Indeed, GWASs of longevity and survival in the general population have identified *APOE* as the strongest genetic factor, with the same ε4 (rs429358) allele associated with increased mortality^26^ and found less frequently in long-living individuals.^27^ Thus, our finding that *APOE* is a risk factor for mortality in PD patients may not be specific to PD.

We also analysed AD GRSs excluding *APOE.* This showed that non-*APOE* AD genetic risk influences mortality in PD, although to a much weaker extent than the *APOE* locus itself.

### TBXAS1

*TBXAS1* encodes Thromboxane A Synthase 1, which catalyses the conversion of prostaglandin H2 to thromboxane A2, which acts as a platelet aggregator and vasoconstrictor. Variants in *TBXAS1* have previously been associated with coronary artery disease in a number of GWASs.^28–31^ We showed that a locus in *TBXAS1* was associated with all-cause mortality in PD cohorts. Thus, it is possible that the association between *TBXAS1* and PD mortality may be due to increased coronary artery disease or other non-PD causes.

However, there are a few important factors to consider. Firstly, the locus we identified for mortality in PD cohorts is different from those associated with coronary artery disease, so it may be that different SNPs within this gene have different effects. Secondly, our search of the most recent longevity GWASs^17^ suggests that variation at *TBXAS1* does not influence mortality and longevity in the general population. Thirdly, it is also possible that patients with PD are more susceptible to adverse effects of a SNP than healthy controls, so PD or its treatment increases vulnerability to the SNP effect on coronary artery disease but the cause of death is not able to be classified as ‘PD-related’. This has also been seen in previous non-genetic studies of PD mortality.^32^ Thus future analyses including cause of death are needed to clarify the mechanism through which *TBXAS1* variants influence mortality in PD patients.

### SYT10

We also found evidence that a locus near *SYT10* (synaptotagmin 10) in Chromosome 12 was associated with mortality. This protein is a calcium sensor involved in regulation of calcium-dependent exocytosis, specifically related to peptidergic vesicles.^33^ Variants in this gene have been linked to heart rate and cardiac cycle phenotypes.^34–36^ This locus was not significantly associated with longevity in the general population, although there was nominal association in the most recent longevity GWAS, (beta=-0.008, p=0.003)^17^.

### MORN1

This locus in Chromosome 1 affected progression to H&Y3. *MORN1* encodes the MORN repeat containing 1 protein, which contains the Membrane Occupation and Recognition (MORN) repeat. The function of this protein is not very well understood or studied in humans. In the GWAS Catalogue (https://www.ebi.ac.uk/gwas/), SNPs in *MORN1* have been associated with height, coronary artery disease, and mean corpuscular haemoglobin concentration among others.

### ASNS

This locus in Chromosome 7 was associated with progression to H&Y3. The top SNP is about 10.5kb away from the gene *ASNS* encoding Asparagine synthetase. This protein is expressed in the brain, pancreas and stomach (https://www.proteinatlas.org/ENSG00000070669-ASNS). It converts aspartate and glutamine to asparagine and glutamate,^39^ with glutamate playing a crucial role as a neurotransmitter. Extracellular glutamate can be neurotoxic primarily through activation of N-methyl-D-aspartate (NMDA) receptors.^40^ This excitotoxicity may contribute to neuronal death in neurodegenerative diseases including PD and AD.^40,41^

*ASNS* has been implicated in certain cancers, in particular acute lymphoblastic leukemia, where a lack of ASNS protein expression is targeted with asparaginase therapy. Asparaginase catalyses the hydrolysis of asparagine and glutamine to aspartic acid and glutamate, which causes intracellular asparagine levels to fall and thereby starves the leukaemia cells, leading to cell death and suppressed tumour growth.^39^ *ASNS* has also been implicated in other cancers, including pancreatic cancer, ovarian cancer, gastric, and hepatic cancers,^39,42,43^ though some cancers show overexpression of *ASNS*.^44^

Asparaginase had a neuroprotective effect in a cell model of PD, where cells overexpressing SNCA A53T had pathological alpha-synuclein aggregation and increased glutamine levels.^45^ Treatment with asparaginase depleted glutamine and activated the degradation of alpha-synuclein through autophagy and improved mitochondrial function.^45^

### XPO1

This locus in Chromosome 2 was associated with progression to H&Y3. The lead SNP is an intronic variant in *XPO1*, however it is also an eQTL for *KIAA1841* and *AHSA2* in blood, and *C2orf74* in brain cortex. In addition, colocalization analysis implicated colocalization with eQTLs for *PUS10*, though there was not strong evidence for a shared causal SNP. In addition, there were two missense variants within 500kb in LD with the lead SNP, in the genes *C2orf74* and *CCT4*.

XPO1 (Exportin 1) transports other proteins from the nucleus to the cytoplasm, specifically proteins containing nuclear export signal (NES), which include tumour suppressors.^46,47^ XPO1 is overexpressed in many cancers, including pancreatic, gastric, prostate, and colorectal cancer, and overexpression is associated with more rapid disease progression and poorer outcomes.^46,47^ Several XPO1 inhibitors are being investigated as therapeutics, including selinexor which has been approved by the US Food and Drug Administration (FDA) for multiple myeloma and diffuse large B-cell lymphoma.^47^

XPO1 also transports anti-inflammatory factors and growth regulating proteins. In PD mouse model, the XPO1 inhibitor eltanexor prevented dopaminergic neuron loss and microglial activation and also rescued locomotor incoordination.^48^

C2orf74, chromosome 2 open reading frame 74, has not been extensively studied. It may be involved in autoimmune disorders, where differences in gene expression have been found in celiac disease,^49^ and GWAS hits within C2orf74 for celiac disease, ulcerative colitis, and Crohn’s disease may alter gene expression in thymic tissue.^50^ However, so far it has not been studied in PD.

### PDE5A/USP53

This locus in Chromosome 4 was closest to *PDE5A* but also influenced expression of *USP53*. *PDE5A* encodes Phosphodiasterase 5A, a member of the cyclic nucleotide phosphodiesterase (PDE) family. PDEs are responsible for breaking down the cyclic nucleotides: cyclic adenosine monophosphate (cAMP), and cyclic guanosine monophosphate (cGMP). These are intracellular second messengers involved in many central nervous system processes, such as neurogenesis, apoptosis, and plasticity. Changes in PDE expression and cyclic nucleotide levels may be involved in aging, Alzheimer’s disease^51^, Huntington’s disease, and PD.^52^

PDE5 specifically breaks down cGMP. Studies have shown that PDE inhibitors like zaprinast restored long-term depression (a form of synaptic plasticity) in mice overexpressing *SNCA* A53T^53^, and reduced the severity of dyskinesias and increased cAMP and cGMP levels in a hemiparkinsonian rat model.^54^ However *PDE5* is not very highly expressed in the striatum compared to other PDEs and also relative to other tissues, and other PDEs may be important in PD.^51,52^

The top SNP in this locus was also a significant eQTL for *USP53* in blood. This gene encodes Ubiquitin Specific Peptidase 53, a type of deubiquitinase, which are enzymes that removes ubiquitin from proteins. *USP53* has been linked to hepatocellular carcinoma (a type of liver cancer) and esophageal cancer, where its downregulation was found in tumour tissues from patients.^55–57^ Overexpression of USP53 promoted tumour cell death and reduced tumour growth, suggesting an anti-tumour role in certain cancers.^56,57^

### PD risk variants and candidate variants

In line with previous PD progression GWASs, the majority of the 90 PD risk SNPs were not associated with PD progression.^2,3,5,58^ We showed that one variant, rs35749011, linked to *GBA1* p.E326K (also known as p.E365K) was associated with mortality. Interestingly this variant does not cause Gaucher disease or have a major effect on glucosylceramide levels suggesting a dissociation between glucosylceramide and the role of *GBA1* in PD progression. This is consistent with the recently reported trial data reporting a lack of effect of the glucosylceramide synthase inhibitor venglustat in modifying PD progression.^59^

We were not able to replicate findings for other candidate variants nominated from previous PD progression GWASs. We examined results for rs382940 in *SLC44A1* for progression to H&Y stage 3 or greater,^2^ however this was not associated with progression in any of our results.

We also did not replicate findings for variants in *RIMS2*, *WWOX,* and *TMEM108* which have been reported for PD dementia.^5^ The p-values for these variants were all > 0.3 in our GWASs (Supplementary Table 6), although we did not analyse cognitive impairment or dementia in this study.

We did not find evidence to support *APOE* ε2 and *MAPT* H1 haplotype as factors for mortality. We found some evidence suggesting the PSP mortality SNP, rs2242367,^13^ was also associated with more rapid mortality in PD. This SNP was shown to increase expression of *LRRK2*, possibly through regulation of a long coding RNA, *LINC02555* although there was no colocalisation with *LRRK2*.^13^ This finding could indicate that there is some ‘contamination’ of PSP cases in our PD cohorts, as PSP can be frequently misdiagnosed as PD and we did not have pathological diagnosis data on the majority of cases, or alternatively the regulatory effects of this SNP, which is separate from the *LRRK2* PD risk locus, may influence survival in both PD and PSP.

### Limitations

This study is one of the largest GWASs of PD progression and the first large-scale GWAS of PD mortality. However larger sample sizes and longer follow-up are needed to detect variants with smaller effects or lower allele frequencies. Secondly, more data is needed on postmortem pathological diagnosis to conduct cause of death analyses, as some of the mortality variants may relate to general population mortality rather than having a specific effect on PD progression. However, *APOE* is the only gene identified in our mortality GWAS that overlaps with general population longevity GWAS. Another limitation is the heterogeneity between cohorts and PD case selection. Our cohorts tend to be recruited from specialist clinics and groups of patients, and this may lead to a tendency to recruit more atypical patients, or rapidly progressing patients. More population-based studies are needed to improve generalisability of these results. Several of our cohorts are also non-incident, with a delay between symptom onset and study entry, and this mean that we are not able to capture the most rapidly progressing patients.

In addition, we nominated genes from the top SNPs based on physical proximity and eQTL databases, however additional fine-mapping and annotation is needed to prioritise causal variants and genes for each locus. Finally, the interpretation of GWAS for neurological disease remains limited by the resolution of the effects of genomic variants on gene expression from bulk RNA sequencing studies. Rapidly increasing sample sizes, and the development of single cell resources will enable a more direct interpretation of the relationship between genomic variants and disease biology.

### Conclusions

We conducted two large-scale GWASs of PD progression, including the first GWAS of mortality in PD. We identified six genome-wide significant signals, including *TBXAS1*. We also showed that the genetic factors influencing progression in PD are largely different to those influencing PD risk, emphasising the need for further studies of progression. This work will help us to better understand the biology of PD progression and develop new disease-modifying treatments.

## Data availability

GWAS summary statistics are available for download at: https://tinyurl.com/PDprogressionv2 (DOI: 10.5281/zenodo.8017385).

All code for our analyses is publicly available at https://github.com/manuelatan/PD-survival-GWAS (DOI: 10.5281/zenodo.7923843).

Tracking Parkinson’s data is available through the Tracking Parkinson’s portal: https://www.trackingparkinsons.org.uk/about-1/data/. The Oxford Parkinson’s Disease Centre Discovery Cohort data (https://doi.org/10.1016/j.parkreldis.2013.09.025) are available on request (Michele Hu). The Cambridgeshire Parkinson’s Incidence from GP to Neurologist (CamPaIGN) (https://www.thebarkerwilliamsgraylab.co.uk/parkinsons-disease/current-studies-pd/) data and the Cambridge clinic data are available on request (Caroline Williams-Gray/ Roger Barker). Parkinson’s Progression Markers Initiative (PPMI) data was accessed from the PPMI platform: https://www.ppmi-info.org/access-data-specimens/download-data. Queen Square Brain Bank for Neurological Disorders (QSBB) data are available upon request. UK Biobank data were accessed through the UK Biobank: https://www.ukbiobank.ac.uk/enable-your-research/apply-for-access. Drug Interaction With Genes in Parkinson’s Disease (DIGPD) data (https://clinicaltrials.gov/ct2/show/NCT01564992) are available upon request (Jean-Christophe Corvol). Calypso data are available on request to the study team (Huw Morris). The Trondheim Parkinson’s Disease Study (Trondheim) data are available on request (https://doi.org/10.14802/jmd.21029). The Oslo Parkinson’s Disease data are available on request (Lasse Pihlstrom/ Mathias Toft).

## Abbreviations

FUMA: Functional Mapping and Annotation of GWAS
GWAS: Genome-Wide Association Study
H&Y: Hoehn and Yahr
LD: Linkage Disequilibrium
MDS-UPDRS: Movement Disorder Society Unified Parkinson’s Disease Rating Scale
PD: Parkinson’s Disease
PPMI: Parkinson’s Progression Markers Initiative
SNP: Single Nucleotide Polymorphism
SD: Standard Deviation
SE: Standard Error
UKB: UK Biobank.

## Data Availability

GWAS summary statistics are available for download at: https://tinyurl.com/PDprogressionv2 (DOI: 10.5281/zenodo.8017385).
All code for our analyses is publicly available at https://github.com/manuelatan/PD-survival-GWAS (DOI: 10.5281/zenodo.7923843).
Tracking Parkinson's data is available through the Tracking Parkinson's portal: https://www.trackingparkinsons.org.uk/about-1/data/. The Oxford Parkinson's Disease Centre Discovery Cohort data (https://doi.org/10.1016/j.parkreldis.2013.09.025) are available on request (Michele Hu). The Cambridgeshire Parkinson's Incidence from GP to Neurologist (CamPaIGN) (https://www.thebarkerwilliamsgraylab.co.uk/parkinsons-disease/current-studies-pd/) data and the Cambridge clinic data are available on request (Caroline Williams-Gray/ Roger Barker). Parkinson's Progression Markers Initiative (PPMI) data was accessed from the PPMI platform: https://www.ppmi-info.org/access-data-specimens/download-data. Queen Square Brain Bank for Neurological Disorders (QSBB) data are available upon request. UK Biobank data were accessed through the UK Biobank: https://www.ukbiobank.ac.uk/enable-your-research/apply-for-access. Drug Interaction With Genes in Parkinson's Disease (DIGPD) data (https://clinicaltrials.gov/ct2/show/NCT01564992) are available upon request (Jean-Christophe Corvol). Calypso data are available on request to the study team (Huw Morris). The Trondheim Parkinson's Disease Study (Trondheim) data are available on request (https://doi.org/10.14802/jmd.21029). The Oslo Parkinson's Disease data are available on request (Lasse Pihlstrom/ Mathias Toft).

https://github.com/manuelatan/PD-survival-GWAS

## Acknowledgements

This research was funded in whole or in part by Aligning Science Across Parkinson’s [Grant number: ASAP-000478] through the Michael J. Fox Foundation for Parkinson’s Research (MJFF). For the purpose of open access, the author has applied a CC BY public copyright license to all Author Accepted Manuscripts arising from this submission.

This research has been conducted using the UK Biobank Resource under Application Number 46450.

Data used in the preparation of this article were obtained from the Parkinson’s Progression Markers Initiative (PPMI) database (https://www.ppmi-info.org/accessdata-specimens/download-data). For up-to-date information on the study, visit www.ppmi-info.org. PPMI – a public-private partnership – is funded by The Michael J. Fox Foundation for Parkinson’s Research and funding partners, including 4D Pharma, AbbVie Inc., AcureX Therapeutics, Allergan, Amathus Therapeutics, Aligning Science Across Parkinson’s (ASAP), Avid Radiopharmaceuticals, Bial Biotech, Biogen, BioLegend, BlueRock Therapeutics, Bristol Myers Squibb, Calico Life Sciences LLC, Celgene Corporation, DaCapo Brainscience, Denali Therapeutics, The Edmond J. Safra Foundation, Eli Lilly and Company, Gain Therapeutics, GE Healthcare, GlaxoSmithKline, Golub Capital, Handl Therapeutics, Insitro, Janssen Pharmaceuticals, Lundbeck, Merck & Co., Inc., Meso Scale Diagnostics, LLC, Neurocrine Biosciences, Pfizer Inc., Piramal Imaging, Prevail Therapeutics, F. Hoffmann-La Roche Ltd and its affiliated company Genentech Inc., Sanofi Genzyme, Servier, Takeda Pharmaceutical Company, Teva Neuroscience, Inc., UCB, Vanqua Bio, Verily Life Sciences, Voyager Therapeutics, Inc., and Yumanity Therapeutics, Inc. (https://www.ppmi-info.org/about-ppmi/who-we-are/study-sponsors).

## Ethics

Tracking Parkinson’s: West of Scotland Research Ethics Service (WoSRES) Research Ethics Committee gave ethical approval for this study (ref 11/AL/0163). Oxford Discovery: NRES Committee, South Central Oxford A Research Ethics Committee gave ethical approval for this study (ref 16/SC/0108). CamPaIGN: Cambridge Research Ethics Committee gave ethical approval for this study. Cambridge PD Research Clinic: Cambridge Research Ethics Committee gave ethical approval for this study. PPMI: The Research Subjects Review Board at the University of Rochester approved the PPMI study protocol. UK Biobank: UK Biobank has approval from the North West Multi-centre Research Ethics Committee (MREC) as a Research Tissue Bank (RTB). QSBB: London Central Research Ethics Committee gave ethical approval for this research tissue bank (ref 18/LO/0721). DIGPD: the Ethical committee Ile-De-France VI gave ethical approval for this study (ID project: 2009-A00109-48). Calypso: Wales Research Ethics Committee 3 gave ethical approval for this study. Trondheim: the Ethics Committee of Central Norway gave ethical approval for this study (ref 2011/1137). Oslo: the Regional Committee for Medical Research Ethics in South-East Norway gave ethical approval for this study.

