## Supplementary Materials for "Genome-wide determinants of mortality and motor progression in Parkinson’s disease"

### Cohorts

We studied 12 cohorts from Europe and America: Tracking Parkinson's[1], Oxford Discovery[2], Parkinson's Progression Markers Initiative (PPMI)[3], Queen Square Brain Bank (QSBB) pathologically-confirmed PD cases, Calypso[4], UK Biobank incident cases and UK Biobank prevalent cases (see below for details), Cambridgeshire Parkinson's Incidence from GP to Neurologist (CamPaIGN)[5,6], the Cambridge PD Research Clinic cohort, Drug Interaction With Genes in Parkinson's Disease (DIGPD)[7], the Trondheim Parkinson's Disease study (Trondheim)[8], and the Oslo Parkinson's Disease study (Oslo)[9]. Cohorts were excluded if less than 20 individuals met the outcome of interest during the follow-up period, or < 5% of the total cohort size. Small numbers can produce unreliable effect size estimates and extremely wide confidence intervals. All cohorts were included for analysis of mortality, with the exception of the PPMI study as not enough patients met the outcome. For the analysis of other clinical outcomes, not all cohorts had available clinical assessments. For Hoehn and Yahr stage, we analysed data from Tracking Parkinson's, Oxford Discovery, PPMI, DIGPD, and Oslo.

If data was available within a cohort, participants who were known to be re-diagnosed with a non-PD condition were excluded from analyses.

### UK Biobank

PD cases were identified from UK Biobank from hospital episode statistics (HES) with an ICD10 code (G20 for PD) in either the primary or secondary position. PD patients were also identified from self-report and death records. UK Biobank data was downloaded on 13/06/2020 (application 46450). PD patients were classified as either prevalent, incident, or undefined, following the 'Definitions of Parkinson's Disease and the major causes of Parkinsonism: UK Biobank Phase 1 Outcomes Adjudication' document (version 1.0, March 2018; http://biobank.ctsu.ox.ac.uk/showcase/showcase/docs/alg_outcome_pdp.pdf). Briefly, prevalent cases were defined as PD patients who had the first PD ICD code date prior to the baseline assessment, or self-reported PD at the baseline assessment. Incident cases were defined as patients with PD detected by HES with the PD ICD code date after the date of baseline assessment. Patients with PD coded in any position in the death register records, but did not have PD in the HES records at any point were also defined as incident cases but these patients were excluded from our analysis. Patients who did not self-report

PD at baseline but a later follow-up visit, and who did not have PD in any HES records or death register records were classified as ‘undefined’. These patients were also excluded from analysis. In our study, we analysed prevalent and incident PD cases separately.

The date of PD diagnosis was defined according to UK Biobank guidelines, using the earliest date of the PD code from HES or self-report. This date of diagnosis was used as a proxy for PD onset in analysis.

Version 2 of the UK Biobank genotype data was used. Quality control and imputation as described below was performed only in the subset of PD cases in the UK Biobank, rather than existing data on the whole cohort.

**Genotyping quality control and imputation**

Genotyping, quality control, and imputation was performed in each cohort separately but following the same steps. Standard quality control procedures were performed in PLINK v1.9 to remove low quality variants, samples, related individuals, and ancestry outliers. Briefly, individuals with low overall genotyping rates (< 98%), related individuals (Identity-By-Descent PIHAT > 0.1), and heterozygosity outliers (> 2 standard deviations away from the mean) were removed. Individuals whose clinically reported biological sex did not match the genetically determined sex were also removed.

To remove ancestry outliers, Principal Components Analysis (PCA) was conducted on a linkage-disequilibrium (LD) pruned set of variants (removing SNPs with an $r^2 > 0.05$ in a 50kb sliding window shifting 5 SNPs at a time) after merging with European (CEU) samples from the HapMap 3 reference panel. Individuals who were more than 6 standard deviations away from the mean of any of the first 10 principal components were removed.

Variants were removed if they had a low genotyping rate (< 99%), Hardy-Weinberg Equilibrium p-value $< 1 \times 10^{-5}$, or minor allele frequency < 1%.

Following quality control, genotypes from each cohort were imputed separately using the Michigan Imputation Server. All cohorts were imputed to the Haplotype Reference Consortium panel (r1.1). Only variants with high imputation quality scores (INFO/R2 > 0.8) were retained for analysis, and imputation dosages were converted into hard call genotypes.

Related and duplicated individuals across cohorts were identified by merging individual level genotype data. One individual from each pair of related individuals was removed (PIHAT > 0.1).

### Statistical analysis

We assessed the following clinical outcomes: mortality, and Hoehn and Yahr stage 3 or greater (when postural instability is present).

The time to event was taken as the first visit where the outcome was met. Individuals who were missing data at all timepoints for the assessment were excluded (e.g. if Hoehn and Yahr stage data was missing at all visits, that patient was excluded from the analysis of progression to Hoehn and Yahr stage 3+).

Progression to each clinical milestone from PD onset was assessed using Cox proportional hazard models, adjusting for age at onset, gender, and the first 5 genetic principal components to adjust for population stratification. For mortality, PD onset was used as the starting timepoint. For Hoehn and Yahr stage 3 or greater, the starting timepoint was set as study entry/baseline visit. Analysis was performed in R using the *survival* package.

### Meta-analysis and visualization

Meta-analysis was performed in METAL, using an inverse variance weighted fixed effects model. GWASs with a genomic inflation factor above 1.2 were excluded from the meta-analysis. Genomic control correction was used to adjust the overall alpha error. After meta-analysis, only SNPs that were present in > 1,000 individuals were included in the final results. SNPs with heterogeneous effects across cohorts were also excluded (p-value < 0.05 for Cochran's Q-test for heterogeneity, and/or I squared > 80). Variants with MAF variability greater than 15% across the cohorts were also excluded. The null hypothesis was tested with the standard GWAS significance level of $5 \times 10^{-8}$.

Results were uploaded to Functional Mapping and Annotation of GWAS (FUMA; https://fuma.ctglab.nl/)[10] to annotate, prioritise, and visualize GWAS results. Standard settings were used in FUMA, with the exception of a higher maximum p-value of to identify lead SNPs ($5 \times 10^{-5}$) so that we could report nominal associations. eQTL gene mapping using all tissue types was also used, in addition to positional mapping. Gene and gene-set analysis was performed with MAGMA within FUMA. Forest plots were generated in R v3.6 using the *forestplot* package.

### GCTA-COJO

Genome-wide Complex Trait Analysis conditional and joint analysis (GCTA-COJO version 1.94.1, https://yanglab.westlake.edu.cn/software/gcta/#COJO) was used to identify if there were multiple independent SNPs within the same locus[11,12]. It performs a stepwise model selection procedure to select independently associated SNPs[11,12].

GCTA-COJO requires a reference sample to estimate LD correlations between SNPs. For our reference sample, we used whole genome sequencing data from the Accelerating Medicines Partnership Parkinson's disease (AMP-PD), including both PD cases and healthy controls. Standard quality control filters were applied to the AMP-PD data, as described previously[13] and outlined here. Samples were removed if they had a call rate < 98%, excess heterozygosity (> 2 SDs from the mean heterozygosity rate), mismatching clinical sex and genetically determined sex from X chromosome heterogeneity, or if they were from related individuals (Identity-By-Descent PIHAT > 0.125). Variants were excluded if they had missingness > 5%, minor allele frequency < 1%, or Hardy-Weinberg Equilibrium p-value < $1x10^{-5}$. To remove ancestry outliers, PCA was conducted on a LD-pruned set of variants after merging with CEU+TSI samples from the HapMap 3 reference panel. Individuals who were more than 6 standard deviations away from the mean of any of the first 10 principal components were removed. After all quality control filters had been applied, 9,422 individuals were remaining in the AMP-PD dataset.

AMP-PD data was in genome build hg38 and lifted over to hg19/GRCh37 genome build using liftOver (RRID:SCR_018160; https://genome.sph.umich.edu/wiki/LiftOver) to match the build of the GWAS summary statistics.

### Colocalization

We performed colocalization analyses using *Coloc* version 5.1 (https://chr1swallace.github.io/coloc/index.html).[14] We also used the package *colochelpR* to help prepare datasets for use in *coloc* (https://github.com/RHReynolds/colochelpR, DOI: 10.5281/zenodo.5011869).[15] We used cis-eQTLs in blood from eQTLGen (https://www.eqtlgen.org/cis-eqtls.html),[16] and cis-eQTLs in brain from PsychENCODE (http://resource.psychencode.org/).[17] Both datasets were downloaded on 22/03/2022.

We followed the same method as in Krohn et al.[18,19] for colocalization analysis (https://github.com/RHReynolds/RBD-GWAS-analysis/). For each locus, we examined all genes with 1Mb of a significant locus in the PD mortality GWAS ($p < 5 \times 10^{-8}$). Coloc was run using default priors. These are the prior probabilities that any random SNP in the region is associated with trait 1 or trait 2, $p_1=10^{-4}$ and $p_2=10^{-4}$. We used a threshold of $p_{12}=5 \times 10^{-6}$ for the $p_{12}$ prior, which is the probability that a SNP in the region is associated with both traits. Loci with a posterior probability of hypothesis 4 (PP.H4) ≥ 0.75 were considered colocalized due to a single shared causal variant, rather than two distinct causal variants (PP.H3).

**Fine-mapping with Probabilistic Annotation INtegraTOR (PAINTOR)**

The top 10 independent loci from each GWAS (Table 2 and Table 3) were selected for statistical fine-mapping with PAINTOR v3.0[20–22]. We followed the recommended pipeline at the PAINTOR v3.0 wiki (https://github.com/gkichaev/PAINTOR_V3.0/wiki) and which has been used in other GWASs[23]. Firstly, a region of 50kb around the most significant SNP was selected (+/- 25kb). Z-scores were calculated from the GWAS summary statistics beta effect size and p-value:

$$Z = \text{sign(Effect Size)} \times \Phi^{-1}(p/2)$$

where $\Phi^{-1}$ is the inverse cumulative distribution function of the normal distribution. Z-scores were calculated in R using the *zsc* function from the *dotgen* package. Secondly, linkage disequilibrium was computed from the 1000 Genomes (Phase 3) reference data for each of the loci. Thirdly, an annotation matrix was created for each locus using all the annotations in the annotation library provided by PAINTOR. Finally, PAINTOR was run on all loci together, on each annotation independently. This was done using the default settings in PAINTOR, which performs approximate inference and enumeration under the assumption of 2 causal variants per locus. The fine-mapped variants with posterior probability > 0.9 are reported in Supplementary Tables 7 and 8. However, it is important to consider that statistical fine-mapping methods are limited and may not necessarily identify causal SNPs in all GWAS loci; functional validation is required to confirm candidate variants.

**PD risk SNPs and GRS**

We also performed candidate variant analysis of the 90 PD risk SNPs from case-control GWAS,[24] and the PD genetic risk score (GRS). The GRS is a cumulative risk score for each individual created from the sum of the genome-wide significant PD risk alleles weighted by effect size. The GRS was created in PLINK v1.9 using the 90 genome-wide significant loci from Nalls et al.[24] The standardised risk score was analysed in each cohort for each outcome using Cox proportional hazard models, adjusting for age at onset, sex, and the PC1-PC5. We created and tested the GRS in each cohort separately and then meta-analysed results using random-effects meta-analysis in R using the package *meta*.

**Candidate variant analysis**

We also examined associations for other candidate variants that have been implicated in PD progression. Previous large-scale genome-wide association studies have identified variants in *SLC44A1* for progression to Hoehn and Yahr stage 3 or greater,[25] and variants in *RIMS2*, *WWOX,* and *TMEM108* for progression to dementia.[26] We also examined results for rs7412 tagging the *APOE* ε2 allele, rs8070723 tagging the *MAPT* H1 haplotype, and rs2242367 adjacent to the LRRK2 locus, which was associated with survival in Progressive Supranuclear Palsy (PSP).[27] We extracted the results for these 7 SNPs from each of our progression GWAS meta-analysis results. We applied Bonferroni correction to adjust for multiple testing for the number of variants tested, with p-value threshold p = 0.05/7 = 0.007.

**Alzheimer's disease genetic risk**

To determine whether the association results for mortality were specific to *APOE* or more general Alzheimer's disease (AD) genetic risk, we analysed the AD GRS in relation to PD progression. 38 loci passing genome-wide significance from the latest AD GWAS were used to create the AD GRS.[28] The *APOE* region was excluded from the GRS (19:40,000,000-50,000,000).[28] AD GRSs were created in each cohort separately (excluding those that had been excluded in the meta-analysis), standardised, then analysed in a Cox proportional hazard model, adjusting for age at onset, sex, and the PC1-PC5. Results were then meta-analysed across cohorts using a random-effects meta-analysis, using the R package *meta*.

**Longevity GWASs**

To help clarify whether our mortality GWAS results were specific to PD mortality or more general mortality/survival, we searched the most recent longevity GWAS[29] and the GWAS Catalog (https://www.ebi.ac.uk/gwas/). Summary statistics from the Timmers et al.[29] longevity GWAS were downloaded from https://datashare.ed.ac.uk/handle/10283/3599. We searched these summary statistics for the top SNPs and genes (+/- 1 Mb) from our PD mortality GWAS results.

**SOFTWARE**

METAL (version 2011-03-25): http://www.sph.umich.edu/csg/abecasis/metal/
(RRID:SCR_002013).
R (v3·6) https://cran.r-project.org/. R Project for Statistical Computing
(RRID:SCR_001905).
PLINK (v1.9): https://www.cog-genomics.org/plink/ PLINK (RRID:SCR_001757).
LocusZoom: http://locuszoom.org/ (RRID:SCR_009257)
liftOver: https://genome.sph.umich.edu/wiki/LiftOver (RRID:SCR_018160).
GCTA-COJO (v1.94.1): https://yanglab.westlake.edu.cn/software/gcta/#COJO
MAGMA: https://snp-magma.sourceforge.net (RRID:SCR_005757)
PAINTOR (v3.1): https://bogdan.dgsom.ucla.edu/pages/paintor/
FUMA (v1.5.1): https://fuma.ctglab.nl/ (RRID:SCR_017521)

R packages for specific analyses:

- *survSNP:* https://cran.r-project.org/web/packages/survSNP/index.html
- *Coloc* (v5.1): https://cran.r-project.org/web/packages/coloc/, https://chr1swallace.github.io/coloc/index.html
- *colochelpR:* https://github.com/RHReynolds/colochelpR
- *forestplot* (v3.3.1)*:* https://cran.r-project.org/web/packages/forestplot/
- *survival* (RRID:SCR_021137)*:* https://cran.r-project.org/web/packages/survival/index.html
- *meta* (RRID:SCR_019055)*:* https://cran.r-project.org/web/packages/meta/index.html

### PUBLIC DATABASES

GWAS Catalogue: https://www.ebi.ac.uk/gwas
LDpair, LDproxy (version 5.6.2): https://ldlink.nih.gov/
eQTLGen: https://www.eqtlgen.org/
MetaBrain: https://www.metabrain.nl/
GTEx: https://gtexportal.org/
PsychENCODE: http://resource.psychencode.org/
Michigan Imputation Server: https://imputationserver.readthedocs.io/en/latest/
HapMap 3 reference panel:
https://www.sanger.ac.uk/resources/downloads/human/hapmap3.html/
https://ftp.ncbi.nlm.nih.gov/hapmap/
Iwaki 2019 Progression GWAS summary statistics:
https://pdgenetics.shinyapps.io/pdprogmetagwasbrowser/
Timmers 2020 longevity GWAS: www.longevitygenomics.org/downloads

### DIGPD STUDY GROUP

Steering committee: Jean-Christophe Corvol, MD (Pitié-Salpêtrière Hospital, Paris, principal investigator of DIGPD); Alexis Elbaz, MD (CESP, Villejuif, member of the steering committee and PI for statistical analyses), Marie Vidailhet, MD (Pitié-Salpêtrière Hospital, Paris, member of the steering committee), Alexis Brice, MD (Pitié-Salpêtrière Hospital, Paris, member of the steering committee and PI for genetic analysis) ; Statistical analyses: Fanny Artaud, PhD (CESP, Villejuif, statistician); Principal investigators for sites: Frédéric Bourdain, MD (CH Foch, Suresnes, PI for site), Jean-Philippe Brandel, MD (Fondation Rothschild, Paris, PI for site), Pascal Derkinderen, MD (CHU Nantes, PI for site), Franck Durif, MD (CHU Clermont-Ferrand, PI for site), Richard Levy, MD (CHU Saint-Antoine, Paris, PI for site), Fernando Pico, MD (CH Versailles, PI for site), Olivier

Rascol, MD (CHU Toulouse, PI for site); Co-investigators: Cecilia Bonnet, MD (Pitié-Salpêtrière Hospital, Paris, site investigator), Christine Brefel-Courbon, MD (CHU Toulouse, site investigator), Florence Cormier-Dequaire, MD (Pitié-Salpêtrière Hospital, Paris, site investigator), Andreas Hartmann, MD (Pitié-Salpêtrière Hospital, Paris, site investigator), Stephan Klebe, MD (Pitié-Salpêtrière Hospital, Paris, site investigator), Julia Kraemmer, MD (Pitié-Salpêtrière Hospital, site investigator), Lucette Lacomblez (Pitié-Salpêtrière Hospital, Paris, site investigator), Graziella Mangone, MD (Pitié-Salpêtrière Hospital, Paris, site investigator), Ana-Raquel Marques, MD (CHU Clermont Ferrand, site investigator), Valérie Mesnage, MD (CHU Saint Antoine, Paris, site investigator), Julia Muellner (Pitié-Salpêtrière Hospital, Paris, site investigator), Fabienne Ory-Magne, MD (CHU Toulouse, site investigator), Hana You (Pitié-Salpêtrière Hospital, Paris, site investigator); Neuropsychologists: Eve Benchetrit, MS (Pitié-Salpêtrière Hospital, Paris, neuropsychologist), Julie Socha, MS (Pitié-Salpêtrière Hospital, Paris, neuropsychologist), Fanny Pineau, MS (Pitié-Salpêtrière Hospital, Paris, neuropsychologist); Sponsor activities and clinical research assistants: Alain Mallet, PhD (Pitié-Salpêtrière Hospital, Paris, sponsor representative), Coralie Villeret (Hôpital Saint Louis, Paris, Project manager), Merry Mazmanian (Pitié-Salpêtrière Hospital, Paris, project manager), Hakima Manseur (Pitié-Salpêtrière Hospital, Paris, clinical research assistant), Mostafa Hajji (Pitié-Salpêtrière Hospital, Paris, data manager), Benjamin Le Toullec, MS (Pitié-Salpêtrière Hospital, Paris, clinical research assistant).

**AMP PD Acknowledgement**

Data used in the preparation of this article were obtained from the Accelerating Medicine Partnership® (AMP®) Parkinson’s Disease (AMP PD) Knowledge Platform. For up-to-date information on the study, visit https://www.amp-pd.org.

The AMP® PD program is a public-private partnership managed by the Foundation for the National Institutes of Health and funded by the National Institute of Neurological Disorders and Stroke (NINDS) in partnership with the Aligning Science Across Parkinson's (ASAP) initiative; Celgene Corporation, a subsidiary of Bristol-Myers Squibb Company; GlaxoSmithKline plc (GSK); The Michael J. Fox Foundation for Parkinson's Research ; Pfizer Inc.; Sanofi US Services Inc.; and Verily Life Sciences.

ACCELERATING MEDICINES PARTNERSHIP and AMP are registered service marks of the U.S. Department of Health and Human Services.

**AMP PD Cohort Acknowledgements**

Clinical data and biosamples used in preparation of this article were obtained from the (i) Michael J. Fox Foundation for Parkinson's Research (MJFF) and National Institutes of Neurological Disorders and Stroke (NINDS) BioFIND study, (ii) Harvard Biomarkers Study (HBS), (iii) National Institute on Aging (NIA) International Lewy Body Dementia Genetics Consortium Genome Sequencing in Lewy Body Dementia Case-control Cohort (LBD), (iv) MJFF LRRK2 Cohort Consortium (LCC), (v) NINDS Parkinson's Disease Biomarkers Program (PDBP), (vi) MJFF Parkinson's Progression Markers Initiative (PPMI), and (vii) NINDS Study of Isradipine as a Disease-modifying Agent in Subjects With Early Parkinson Disease, Phase 3 (STEADY-PD3) and (viii) the NINDS Study of Urate Elevation in Parkinson's Disease, Phase 3 (SURE-PD3).

BioFIND is sponsored by The Michael J. Fox Foundation for Parkinson's Research (MJFF) with support from the National Institute for Neurological Disorders and Stroke (NINDS). The BioFIND Investigators have not participated in reviewing the data analysis or content of the manuscript. For up-to-date information on the study, visit michaeljfox.org/news/biofind.

Genome sequence data for the Lewy body dementia case-control cohort were generated at the Intramural Research Program of the U.S. National Institutes of Health. The study was supported in part by the National Institute on Aging (program #: 1ZIAAG000935) and the National Institute of Neurological Disorders and Stroke (program #: 1ZIANS003154)." "The Harvard Biomarker Study (HBS) is a collaboration of HBS investigators [full list of HBS investigators found at https://www.bwhparkinsoncenter.org/biobank/ and funded through philanthropy and NIH and Non-NIH funding sources. The HBS Investigators have not participated in reviewing the data analysis or content of the manuscript.

Data used in preparation of this article were obtained from The Michael J. Fox Foundation sponsored LRRK2 Cohort Consortium (LCC). The LCC Investigators have not participated in reviewing the data analysis or content of the manuscript. For up-to-date information on the study, visit https://www.michaeljfox.org/biospecimens).

PPMI is sponsored by The Michael J. Fox Foundation for Parkinson's Research and supported by a consortium of scientific partners: [list the full names of all of the PPMI funding partners found at https://www.ppmi-info.org/about-ppmi/who-we-are/study-sponsors]. The PPMI investigators have not participated in reviewing the data analysis or content of the manuscript. For up-to-date information on the study, visit www.ppmi-info.org.

The Parkinson's Disease Biomarker Program (PDBP) consortium is supported by the National Institute of Neurological Disorders and Stroke (NINDS) at the National Institutes of Health. A full list of PDBP investigators can be found at https://pdbp.ninds.nih.gov/policy. The PDBP investigators have not participated in reviewing the data analysis or content of the manuscript.

The Study of Isradipine as a Disease-modifying Agent in Subjects With Early Parkinson Disease, Phase 3 (STEADY-PD3) is funded by the National Institute of Neurological Disorders and Stroke (NINDS) at the National Institutes of Health with support from The Michael J. Fox Foundation and the Parkinson Study Group. For additional study information, visit https://clinicaltrials.gov/ct2/show/study/NCT02168842. The STEADY-PD3 investigators have not participated in reviewing the data analysis or content of the manuscript.

The Study of Urate Elevation in Parkinson's Disease, Phase 3 (SURE-PD3) is funded by the National Institute of Neurological Disorders and Stroke (NINDS) at the National Institutes of Health with support from The Michael J. Fox Foundation and the Parkinson Study Group. For additional study information, visit https://clinicaltrials.gov/ct2/show/NCT02642393. The SURE-PD3 investigators have not participated in reviewing the data analysis or content of the manuscript.
