## Supplementary Figures for "Genome-wide determinants of mortality and motor progression in Parkinson’s disease"

**Supplementary Figure 1. LocusZoom plot for rs429358 in mortality GWAS in Chromosome 19.**

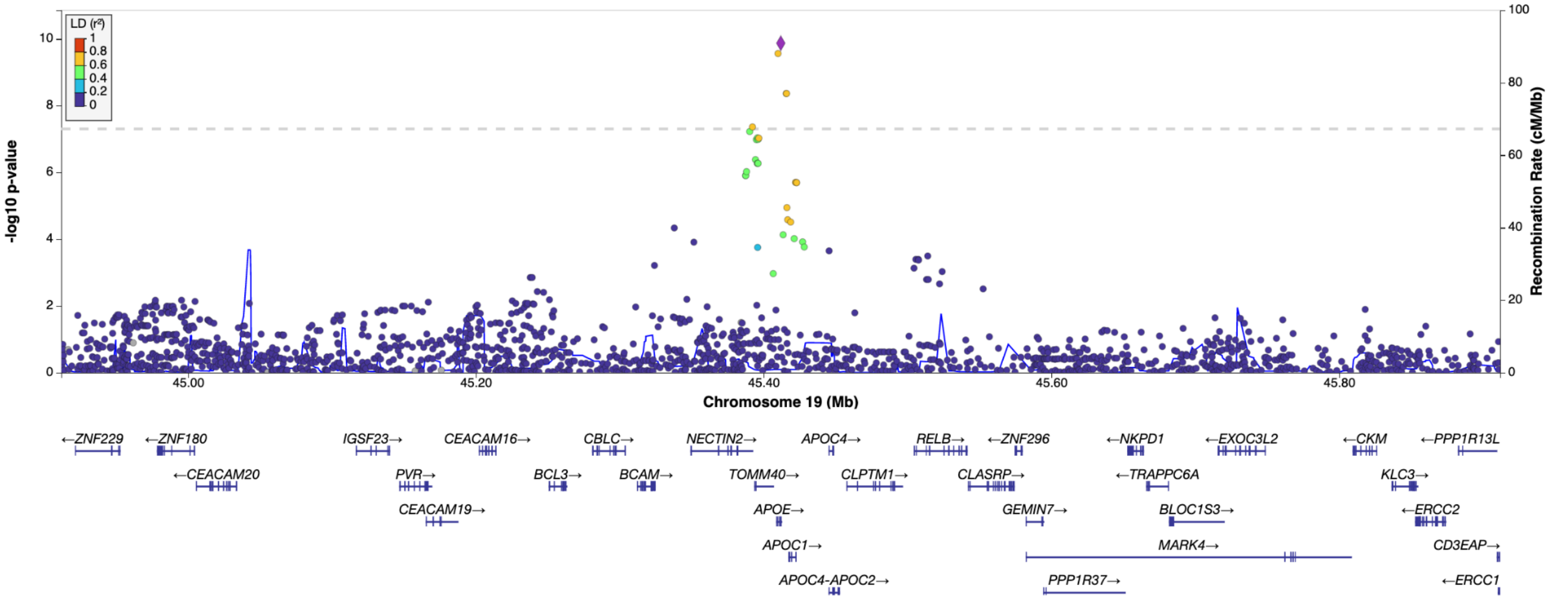


Data is shown in hg37 build. The purple diamond shows the most significantly associated Single Nucleotide Polymorphism (SNP) in the locus. SNPs in linkage disequilibrium (LD) with the index SNP are shown in coloured circles according to the $r^2$, as shown in the legend on the top left. The recombination rate is shown as the blue line with the axis on the right y axis. Genes are shown in the bottom panel with the positions of exons displayed as bars, and transcribed strand indicated with an arrow.

**Supplementary Figure 2. LocusZoom plot for rs4726467 in mortality GWAS in Chromosome 7.**

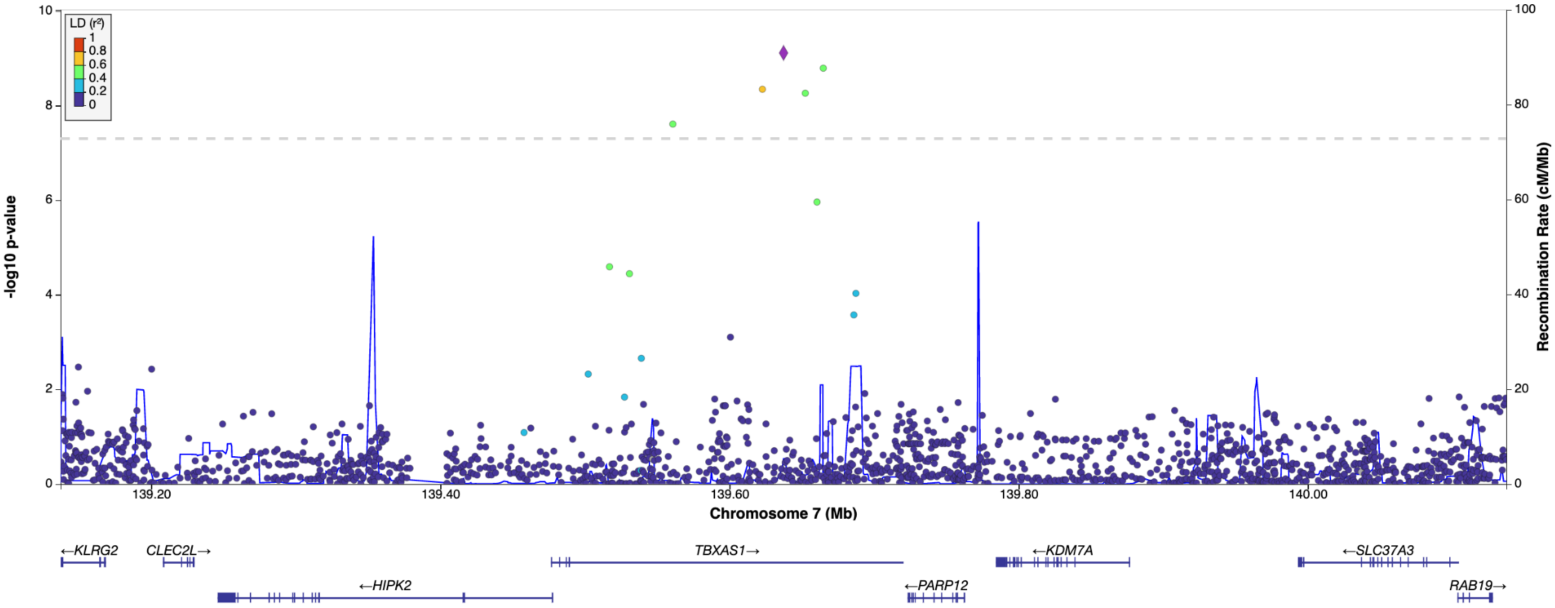


Data is shown in hg37 build. The purple diamond shows the most significantly associated Single Nucleotide Polymorphism (SNP) in the locus. SNPs in linkage disequilibrium (LD) with the index SNP are shown in coloured circles according to the $r^2$, as shown in the legend on the top left. The recombination rate is shown as the blue line with the axis on the right y axis. Genes are shown in the bottom panel with the positions of exons displayed as bars, and transcribed strand indicated with an arrow.

**Supplementary Figure 3. LocusZoom plot for rs10437796 in mortality GWAS in Chromosome 12.**

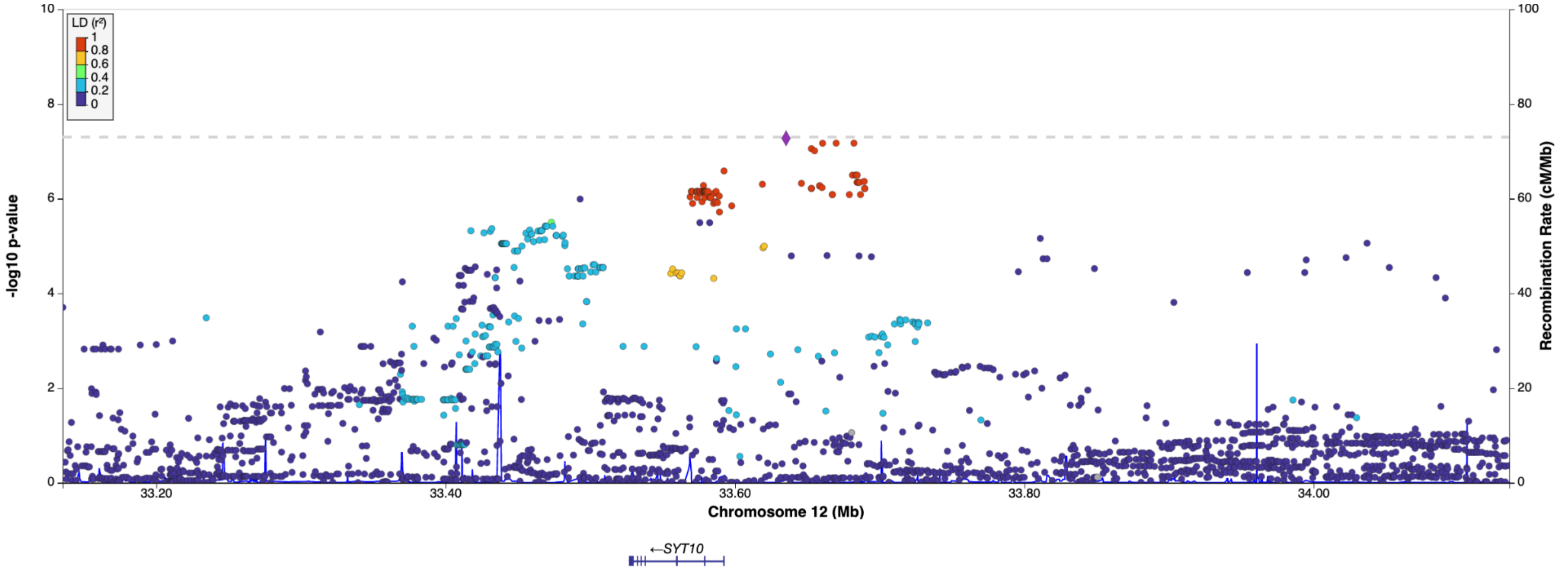


Data is shown in hg37 build. The purple diamond shows the most significantly associated Single Nucleotide Polymorphism (SNP) in the locus. SNPs in linkage disequilibrium (LD) with the index SNP are shown in coloured circles according to the $r^2$, as shown in the legend on the top left. The recombination rate is shown as the blue line with the axis on the right y axis. Genes are shown in the bottom panel with the positions of exons displayed as bars, and transcribed strand indicated with an arrow.

**Supplementary Figure 4. LocusZoom plot for rs115217673 in H&Y3+ GWAS in Chromosome 1.**

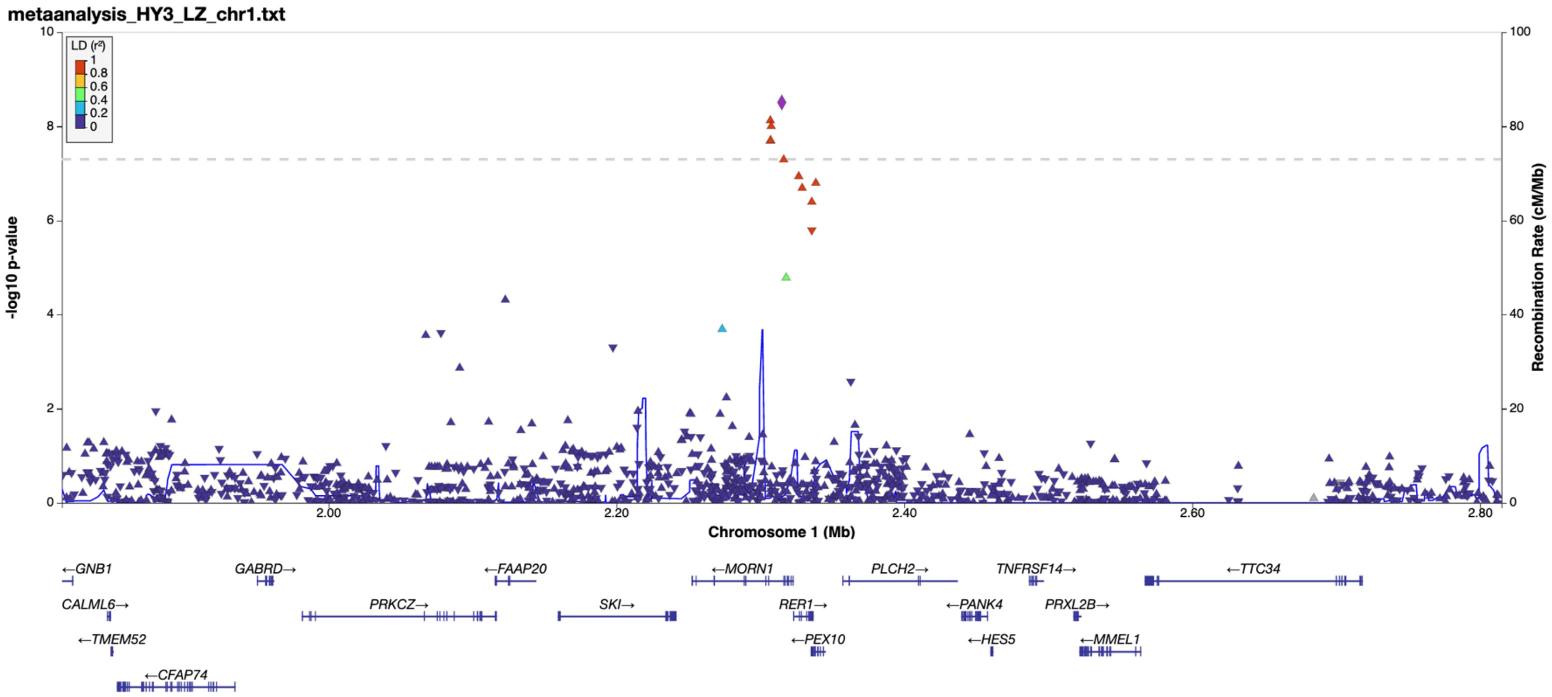


Data is shown in hg37 build. The purple diamond shows the most significantly associated Single Nucleotide Polymorphism (SNP) in the locus. SNPs in linkage disequilibrium (LD) with the index SNP are shown in coloured circles/triangles according to the $r^2$, as shown in the legend on the top left. The recombination rate is shown as the blue line with the axis on the right y axis. Genes are shown in the bottom panel with the positions of exons displayed as bars, and transcribed strand indicated with an arrow.

**Supplementary Figure 5. LocusZoom plot for rs141421624 in H&Y3+ GWAS in Chromosome 2.**

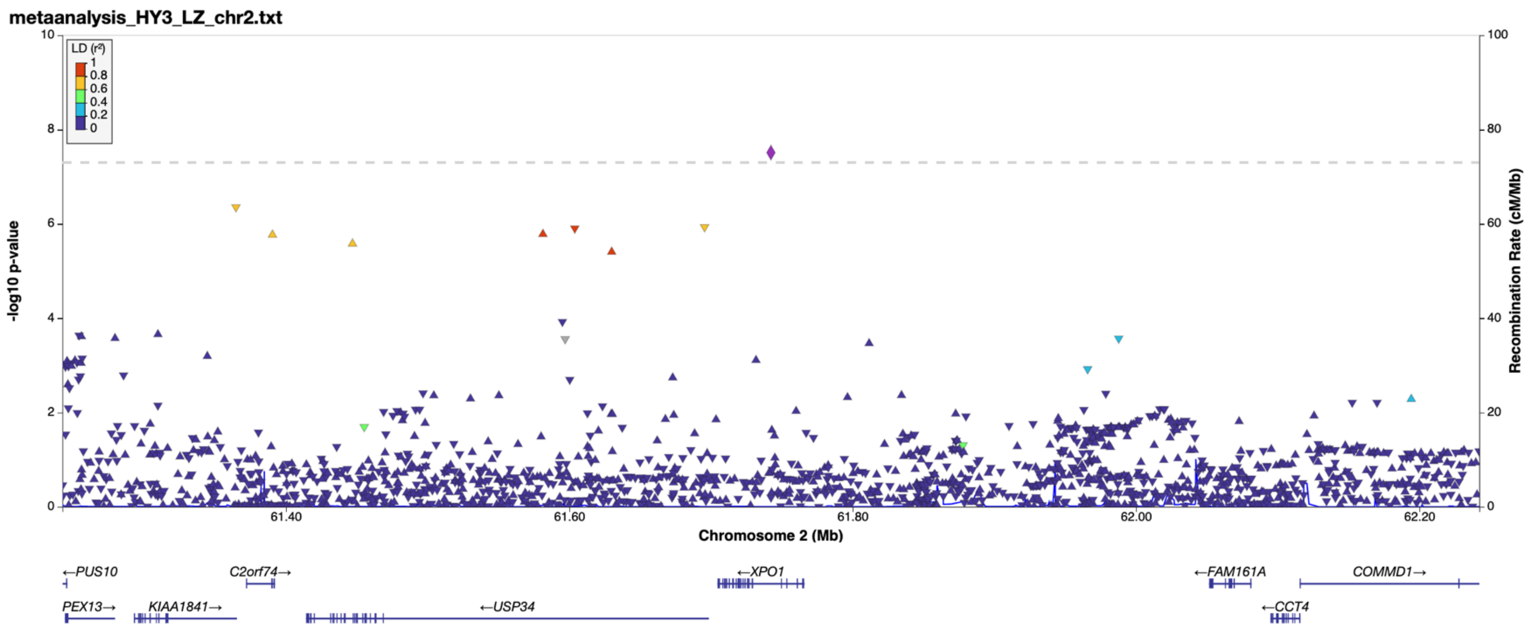

**Supplementary Figure 6. LocusZoom plot for rs113120976 in H&Y3+ GWAS in Chromosome 4.**

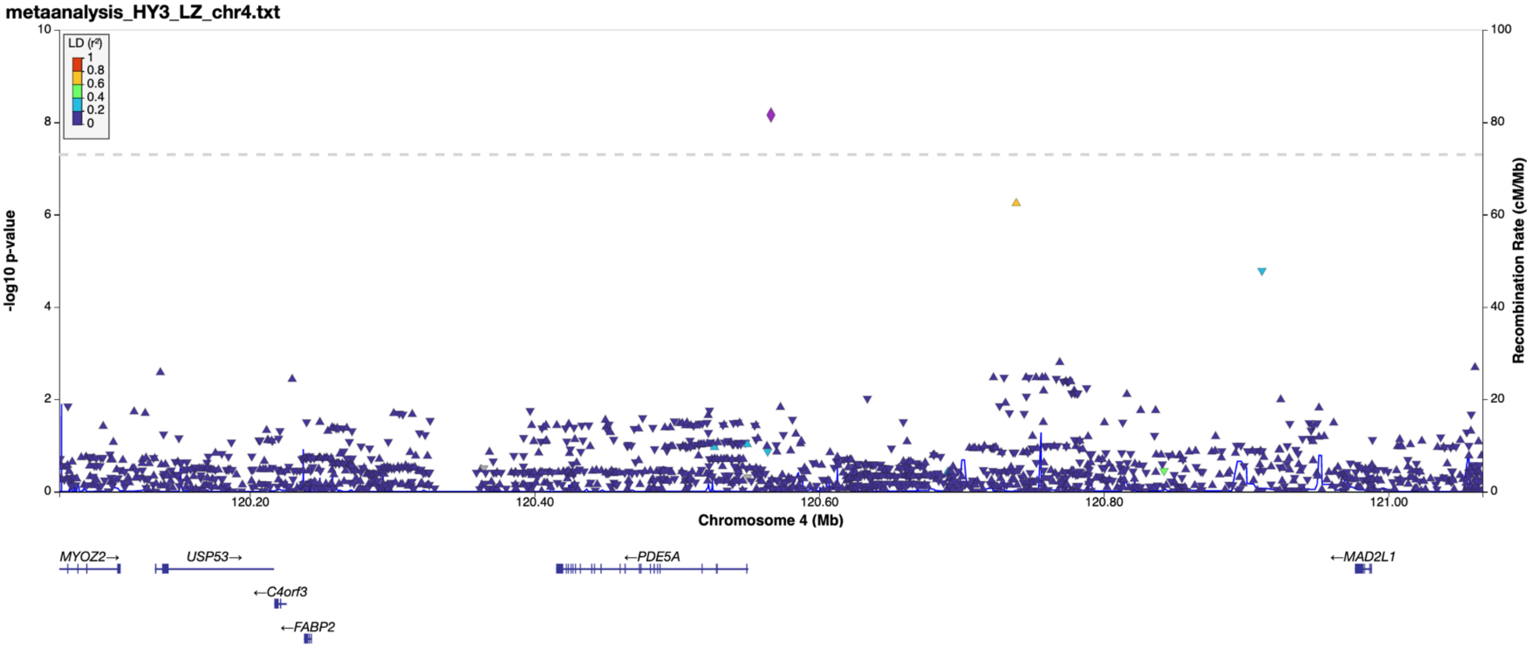


Data is shown in hg37 build. The purple diamond shows the most significantly associated Single Nucleotide Polymorphism (SNP) in the locus. SNPs in linkage disequilibrium (LD) with the index SNP are shown in coloured circles/triangles according to the $r^2$, as shown in the legend on the top left. The recombination rate is shown as the blue line with the axis on the right y axis. Genes are shown in the bottom panel with the positions of exons displayed as bars, and transcribed strand indicated with an arrow.

**Supplementary Figure 7. LocusZoom plot for rs145274312 in H&Y3+ GWAS in Chromosome 7.**

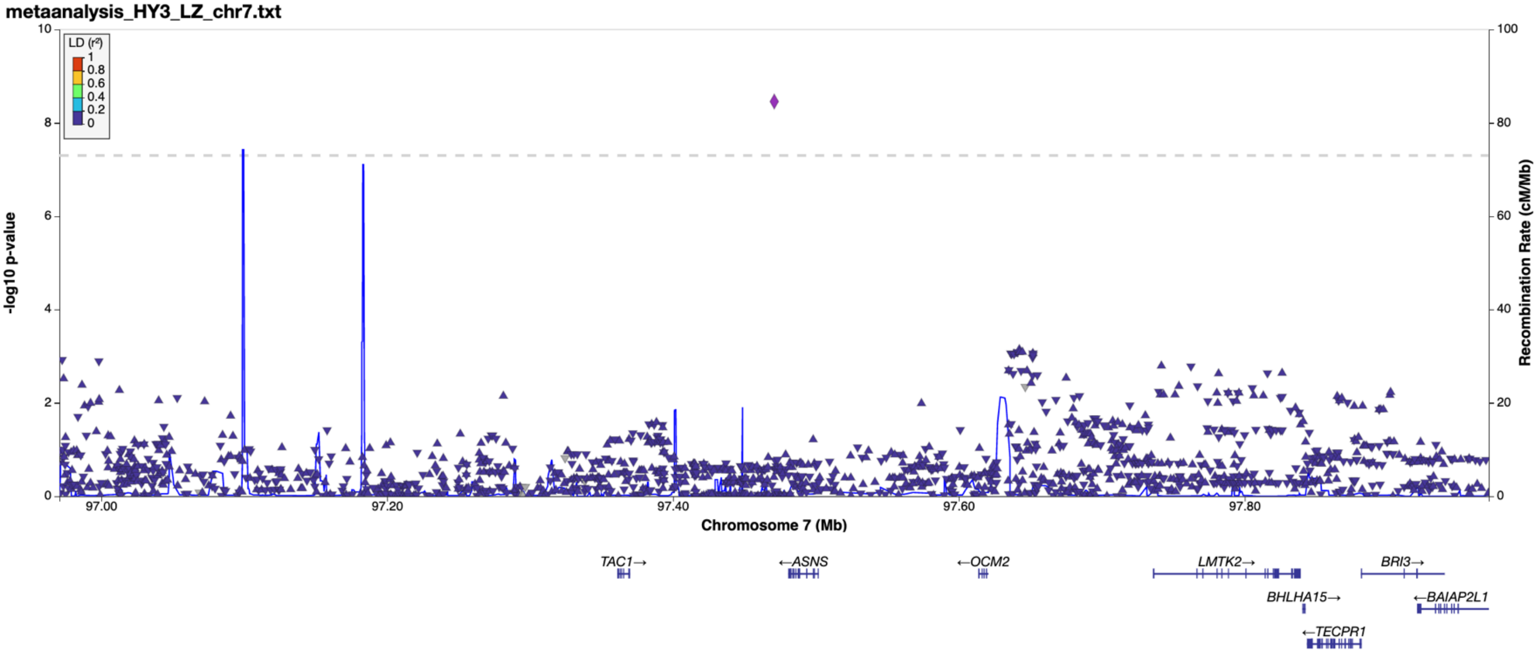


Data is shown in hg37 build. The purple diamond shows the most significantly associated Single Nucleotide Polymorphism (SNP) in the locus. SNPs in linkage disequilibrium (LD) with the index SNP are shown in coloured circles/triangles according to the $r^2$, as shown in the legend on the top left. The recombination rate is shown as the blue line with the axis on the right y axis. Genes are shown in the bottom panel with the positions of exons displayed as bars, and transcribed strand indicated with an arrow.

**Supplementary Figure 8. Regional association plot for eQTL (derived from PsychENCODE) and PD H&Y3 GWAS colocalizations in the region surrounding *PUS10*.**

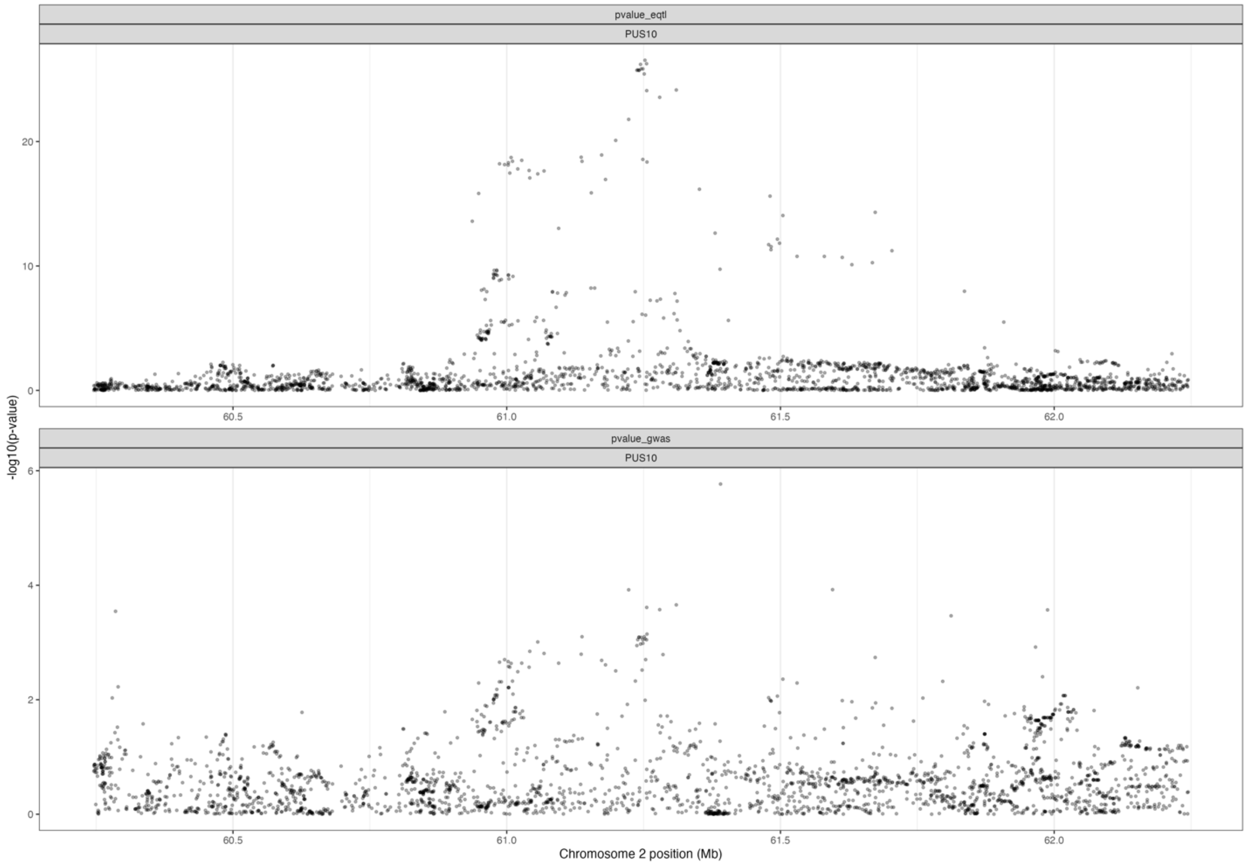


Regional association plot for eQTL and PD H&Y3 GWAS colocalization in the region surrounding *PUS10*. The regional association plot for eQTL are shown in the upper pane, and the PD H&Y3 GWAS association signals are shown in the lower pane. eQTLs were derived from the PsychENCODE, where adult brain tissue from 1,387 individuals was analysed. The PP.H4 (the posterior probability that there is an association in the region with trait 1 and trait 2, with one shared SNP) for *PUS10* was 0.698. The x-axis denotes chromosomal position in hg19, and the y-axis indicates association p-values on a -log10 scale.

**Supplementary Figure 9. Heatmap showing the PD GWAS risk loci from Nalls et al. (2019) that were associated (p < 0.05) with mortality, or H&Y stage 3+. Only variants with at least one association p < 0.05 are shown in the heatmap.**

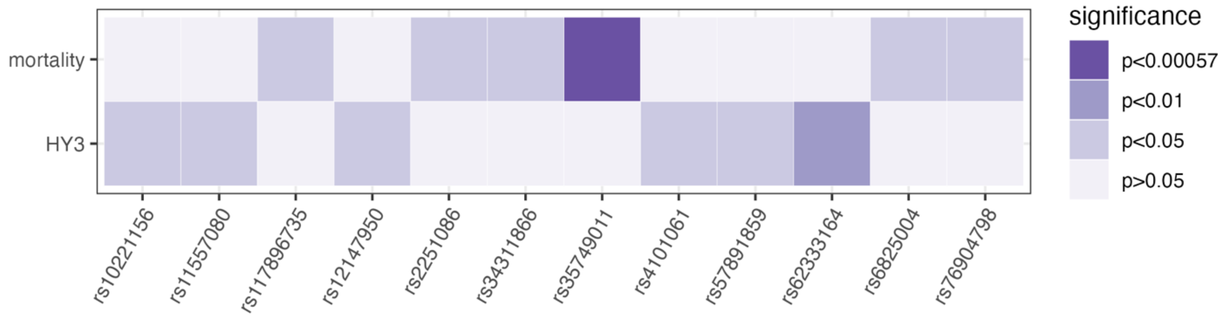

**Supplementary Figure 10. Power calculations for mortality GWAS with N = 5,744 at different effect sizes, allele frequencies, and event rates (shown in faceted plots with grey bars). Calculated using the R package ‘survSNP’.**

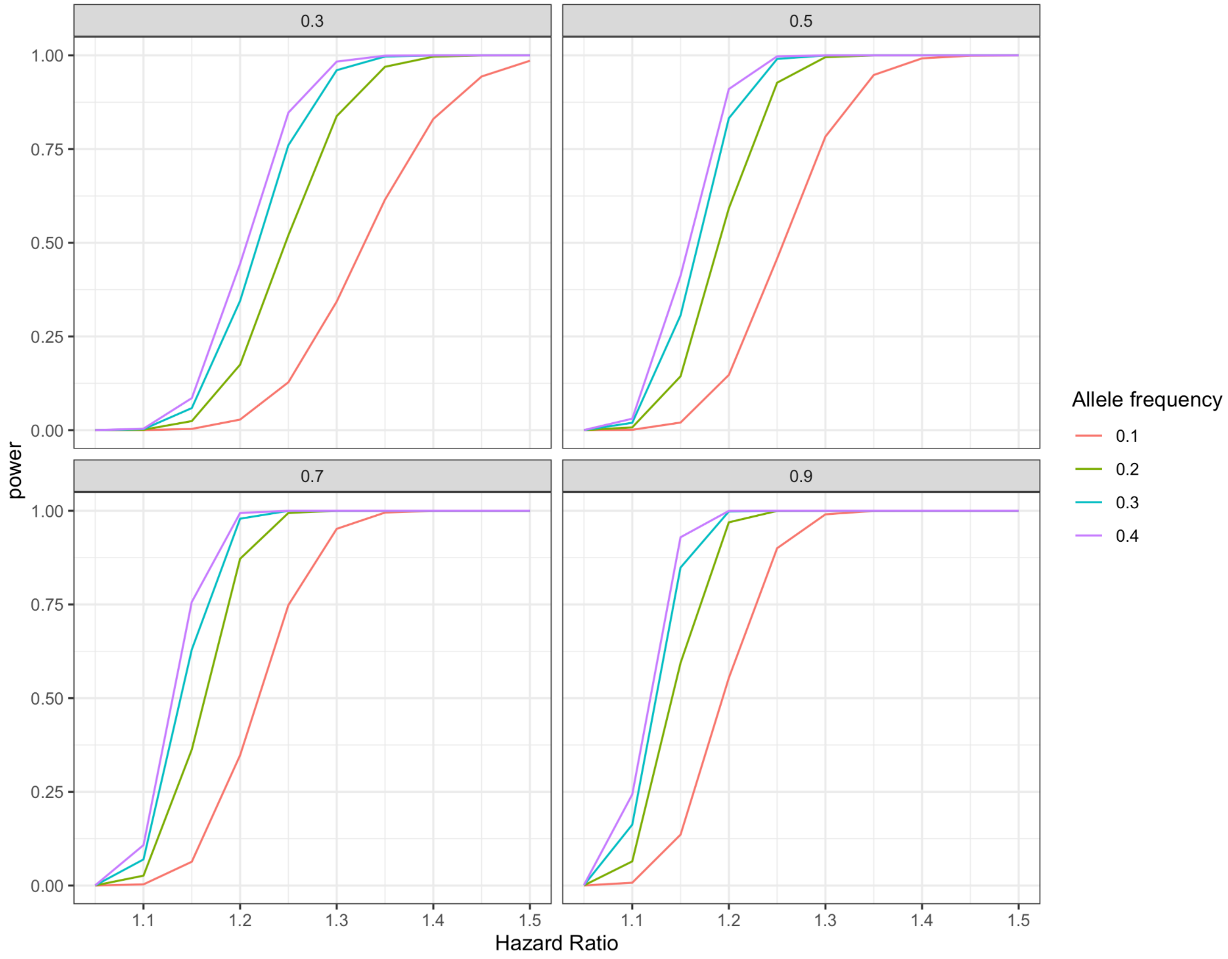

**Supplementary Figure 11. Flowchart showing the fine-mapping pipeline in PAINTOR.**

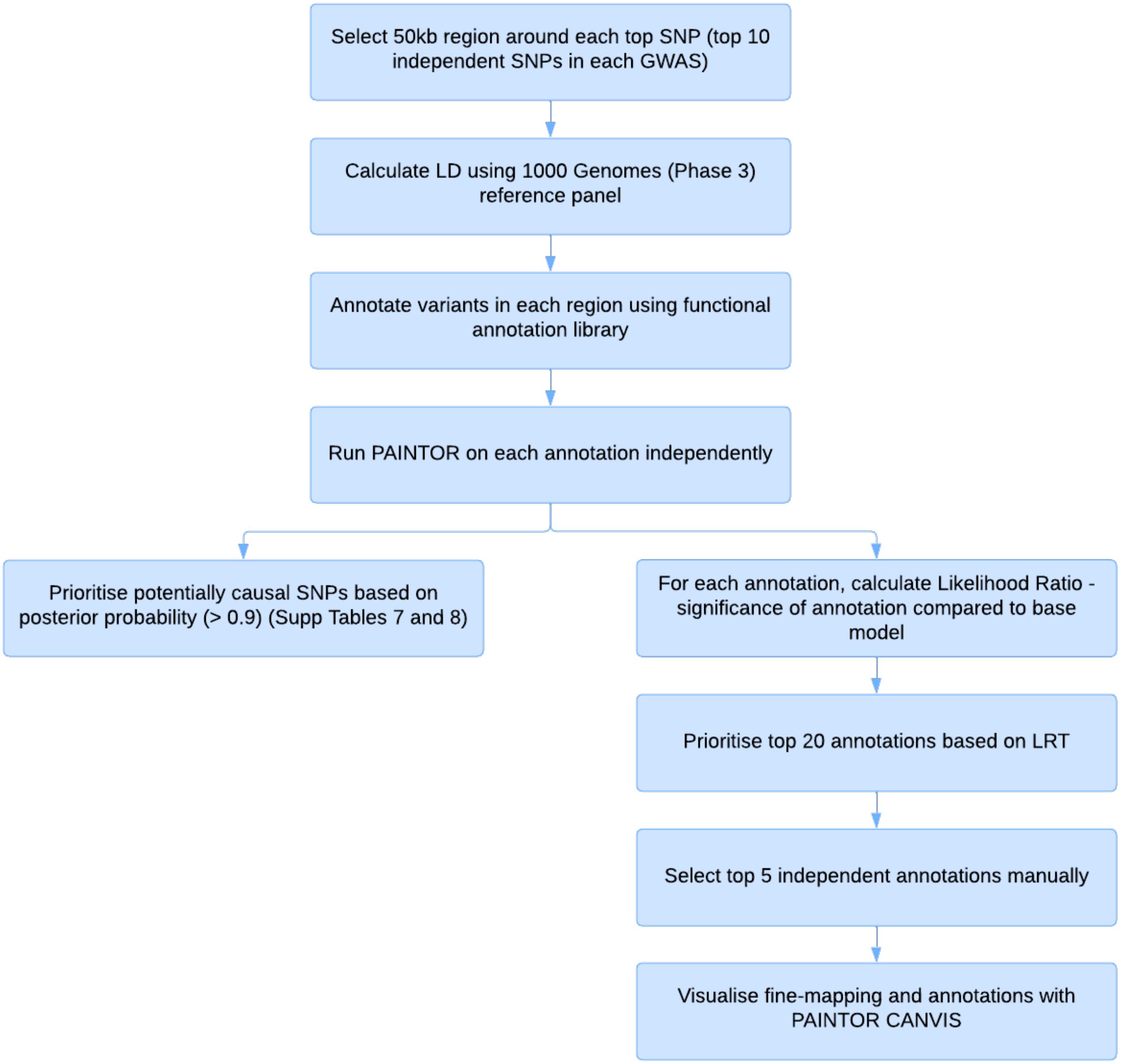
